# Replicating cardiovascular outcome trials of medications used to treat type 2 diabetes using real-world data: A systematic review of observational studies

**DOI:** 10.1101/2024.11.28.24317099

**Authors:** Wanning Wang, Wang-Choi Tang, Michael Webster-Clark, Oriana HY Yu, Kristian B. Filion

**Affiliations:** Department of Epidemiology, Biostatistics and Occupational Health, McGill University, Montreal, Quebec, Canada; Center for Clinical Epidemiology, Lady Davis Institute, Jewish General Hospital, Montreal, Quebec, Canada; Department of Medicine, McGill University, Montreal, Quebec, Canada; Division of Endocrinology and Metabolism, Jewish General Hospital/McGill University, Montreal, Quebec, Canada

**Keywords:** Cardiovascular Outcome Trials, Generalizability, Real-world data, Systematic review

## Abstract

**Background:** Cardiovascular outcome trials (CVOTs) are mandated by the U.S. Food and Drug Administration to assess the cardiovascular safety of new antidiabetic medications before entering the market. However, CVOTs often involve highly selective populations and results may not generalize to real-world settings.

**Methods:** Our study aimed to synthesize observational studies to assess the generalizability of CVOTs to real-world settings. We systematically reviewed observational studies that emulated previous CVOTs for dipeptidyl peptidase-4 (DPP-4) inhibitors, glucagon-like peptide 1 (GLP-1) receptor agonists, and sodium glucose cotransporter-2 (SGLT-2) inhibitors among patients with type 2 diabetes. We searched the MEDLINE, EMBASE and Cochrane databases for observational studies that focused on trial emulation or cross-sectional studies that reported the proportion of real-world patients eligible for completed CVOTs. Two independent reviewers screened articles, extracted data, and assessed study concordance with randomized controlled trial (RCT) results.

**Results:** Nineteen studies were included in our systematic review, including four cohort studies that emulated previous RCTs and 15 cross-sectional studies that evaluated trial eligibility. Results between RCTs and real-world data (RWD) were concordant for all drug classes in finding non-inferiority. The median eligibility percentage ranged from 13% to 31% for SGLT-2 inhibitor trials and 12% to 43% for GLP-1 receptor agonist trials.

**Conclusions:** These results suggest that, while RCTs and RWD are concordant in their estimates, the trials lack representativeness. More research is needed on the replication of CVOTs using RWD to understand how different replication methods may impact findings.

## INTRODUCTION

Randomized controlled trials (RCTs) are the current gold standard for assessing drug efficacy and for evidence-based regulatory decision making. However, due to their strict inclusion and exclusion criteria, trial populations may differ substantially from the real-world population receiving the medication^1^. In addition, there are situations where it may not be suitable to conduct RCTs such as rare diseases or in populations ineligible for RCTs (e.g., children, pregnant women, the elderly). Thus, real world data (RWD) play an important role in the generation of evidence to evaluate the effectiveness and safety of medical interventions. With the adoption of electronic health records, large amounts of RWD on prescription drug use and health outcomes are becoming readily available.

In 2008, the U.S Food and Drug Administration (FDA) issued recommendations for conducting cardiovascular outcome trials (CVOT) to ensure that antidiabetic medications have acceptable cardiovascular risk profiles^2^. More than 13 CVOTs have been conducted on antidiabetic drugs, including dipeptidyl peptidase-4 (DPP-4) inhibitors, glucagon-like peptide 1 (GLP-1) receptor agonists, and sodium glucose cotransporter-2 (SGLT-2) inhibitors. Results from these trials demonstrated cardiovascular safety, with many showing superiority^3^. However, these trials were highly selective and may not be generalizable to patients in real-world settings. For example, many trials required the presence of high cardiovascular risk factor levels to increase the number of events to achieve greater statistical power^4–6^.

With regulatory agencies increasingly considering real-world data (RWD) alongside randomized controlled trial (RCT) evidence in their decision-making, there is a growing need to understand how these two types of evidence complement each other. This is particularly true for antidiabetic medications, which are tested in large CVOTs with strict inclusion criteria. Therefore, we synthesized available information to understand what proportion of patients treated in real-world setting are eligible for CVOTs and compare patient characteristics and estimated treatment effects using RWD and the respective CVOTs via systematic review of observational studies.

## METHODS

This systematic review was conducted following guidelines described in the Cochrane Handbook^7^ and reported following the Preferred Reporting Items for Systematic Reviews and Meta-Analysis (PRISMA)^8^ and Synthesis Without Meta-analysis (SWiM) guidelines^9^ (**Tables S1-S2**). The study protocol was registered in the Open Science Framework platform (<u>10.17605/OSF.IO/G8ERW</u>).

### Search strategy

We searched MEDLINE, EMBASE, and the Cochrane databases from inception to June 5^th^, 2024, to identify studies that used RWD to examine the proportion of patients eligible for CVOTs that emulated these trials. Our predefined search strategy utilized Medical Subject Headings (MeSH) in OVID MEDLINE, EMTREE terms in EMBASE, and keywords in all databases. The search strategy was defined in consultation with a medical librarian and is described in detail in **Tables S3-S5**. Briefly, we searched using the following concepts: observational study, RCTs, major adverse cardiovascular outcomes (MACE), T2DM, emulation/replication/comparability. There were no restrictions on language or geographic location. We also conducted a hand search of references included in studies (backward search), studies that have referenced identified key studies (forward search) and previous reviews not captured by our initial database search. We also hand-searched the grey literature using Google Scholar (first 10 pages).

### Inclusion and exclusion criteria

We included published, peer-reviewed observational studies (e.g., cohort or nested-case controls studies) that compared the risk of cardiovascular outcomes in real-world patients with T2DM aged 18+ years to that observed in CVOTs. To reduce bias from lack of established temporality, cross-sectional studies were only used to examine eligibility and patient characteristics; they were not included in the assessment of clinical outcomes. The interventions of interest were DPP-4 inhibitors, GLP-1 receptor agonists, and SGLT-2 inhibitors. A list of completed CVOTs and their drug class and molecule can be found in **Table S6**. There were no restrictions on comparators; active comparators such as other antidiabetic medications, standard of care, or lifestyle interventions were eligible for inclusion.

We excluded RCTs, meta-analyses, case reports, case series, letters-to-the-editor, editorials, and commentaries. In addition, we excluded conference abstracts as they often present preliminary results and typically do not undergo rigorous peer review. Full texts published in a language other than English were also excluded.

The primary outcome of interest was MACE, which included cardiovascular death, myocardial infraction (MI), and ischemic stroke. Unstable angina was also included if it was reported as part of the MACE definition. Secondary outcomes were the individual components of MACE, hospitalization for heart failure (HHF), and all-cause mortality. We examined the proportion of the study population eligible for a referenced CVOT as reported by the authors. We also compared the reported patient characteristics of the real-world population and the subpopulation eligible for the CVOTs.

### Study selection

Citations generated from the electronic database search were exported to Covidence^10^ and duplicates were removed. Two independent reviewers (WW and WT) screened each study title and abstract for potential inclusion. Any study deemed potentially eligible by either reviewer proceeded to full-text review, where the full-text of each study was evaluated independently by both reviewers. Disagreements were resolved by consensus or if needed, with input from a third author (KBF).

### Data extraction

Two independent reviewers (WW and WT) extracted data using a piloted Microsoft Excel workbook. Disagreements were resolved via consensus or if needed, with the aid of a third author (KBF).

### Assessment of agreement between RCTs and RWD

To assess generalizability of RCT results in RWD studies we used agreement statistics from the DUPLICATE study to compare effect estimates of primary outcomes between the restricted RWD population and the referenced CVOT population^11,12^. *Full statistical significance agreement* was considered to have occurred when the RWD and RCT estimates and 95% confidence intervals (CIs) were on the same side of the null. *Partial significance agreement* was considered to have occurred when the prespecified noninferiority criteria was met, even if the RWD study indicated superiority^12^. Statistical significance agreement was categorized as yes, no, or partial. *Estimate agreement* was considered to have occurred when the effect estimate of the RWD study fell within the 95% CI of the RCT effect estimate. *Standardized difference agreement* was defined by standardized differences |Z| < 1.96.

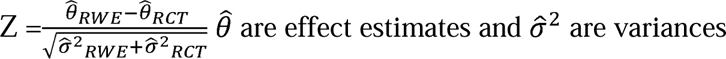

As per the FDA, all the major CVOTs were designed for non-inferiority with an upper CI limit of 1.3^13^. For our statistical significance agreement analysis, we first assessed whether the trial was able to demonstrate non-inferiority. We then evaluated whether the RWD study was able to replicate the non-inferiority finding within the same margin. If both studies demonstrated non-inferiority, then full statistical significance agreement was established. If the trial achieved non-inferiority but the RWD study achieved superiority, partial statistical significance agreement was established. In addition to non-inferiority, if superiority was established in the trial, we assessed if the RWD study also found superiority to achieve full statistical significance agreement.

### Data synthesis

The high degree of clinical heterogeneity among studies and interventions prevented us from conducting a meta-analysis, thus, we reported results following the SWiM reporting guidelines^9^. The main summary measure was the adjusted hazard ratio (HR), as RCTs estimated HRs to assess MACE outcomes. Studies were grouped by drug class and further stratified by RCT emulated. Data were synthesized based on the treatment groups and are presented in tables. Heterogeneity and certainty of evidence were assessed by examining the methods in which the study attempted to replicate the RCT. We captured the exposure definition (intention-to-treat vs on-treatment), active comparators, restriction of the population, follow-up duration, and methods used to reduce confounding.

We conducted several exploratory, post-hoc analyses to examine the potential association of patient characteristics and the proportion of the RWD population eligible for the trial. First, we used scatterplots to identify trends between patient characteristics to the percentage of the population that would have been eligible for their reference CVOT. Second, we calculated standardized mean differences of patient characteristics between the RWD populations and the trial population for select studies based on data completeness.

## RESULTS

### Search results

We identified 6,118 potentially relevant publications (**Figure 1)**. After removing duplicates, 2,389 articles underwent title and abstract review, and 41 underwent full-text review. A total of 19 studies were included in the systematic review^14–32^.

**Figure 1.**
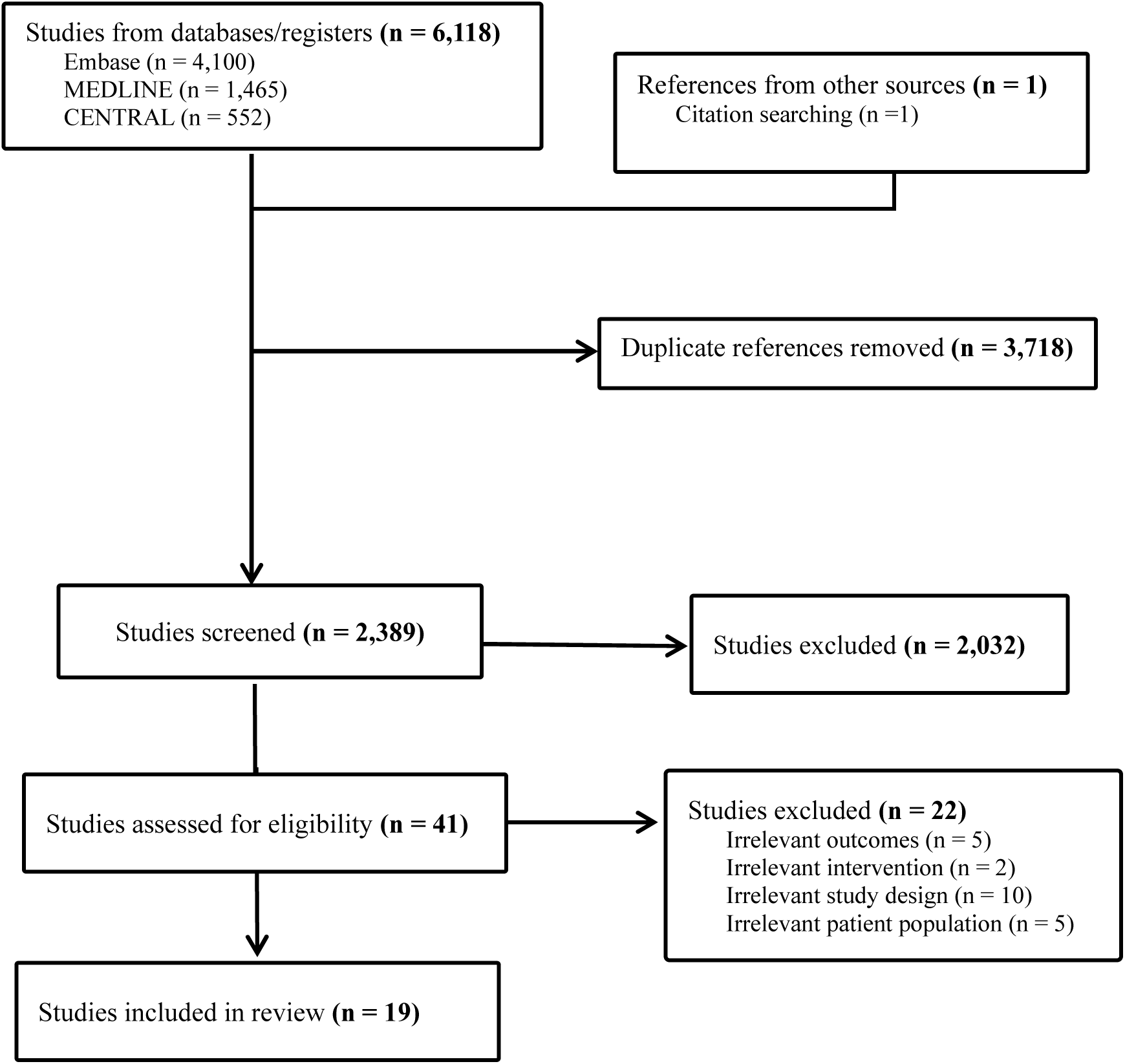
PRISMA flow diagram of study inclusion for systematic review of replication of CVOTs using RWD in patients with T2DM

### Study characteristics

**Table 1** summarizes the characteristics of the included studies stratified by drug class. Four cohort studies^11,21,31,32^ examined ten unique populations for MACE. We identified 15 cross-sectional studies that examined the proportion eligible and patient characteristics^14,16–20,22–30^. The included studies were conducted between 2009 and 2020 in the US (9), Europe (7), and Asia (3). A total of 10 studies assessed GLP-1 receptor agonists^11,16–18,21,23,25,27,30,31^, 12 assessed SGLT-2 inhibitors^11,14,18,20–22,24,26–29,32^, and two assessed DPP-2 inhibitors^11,21^. Within the drug classes, 13 studies evaluated multiple CVOTs (range: 1 to 7 CVOTs; median: 3 per class).

**Table 1.**
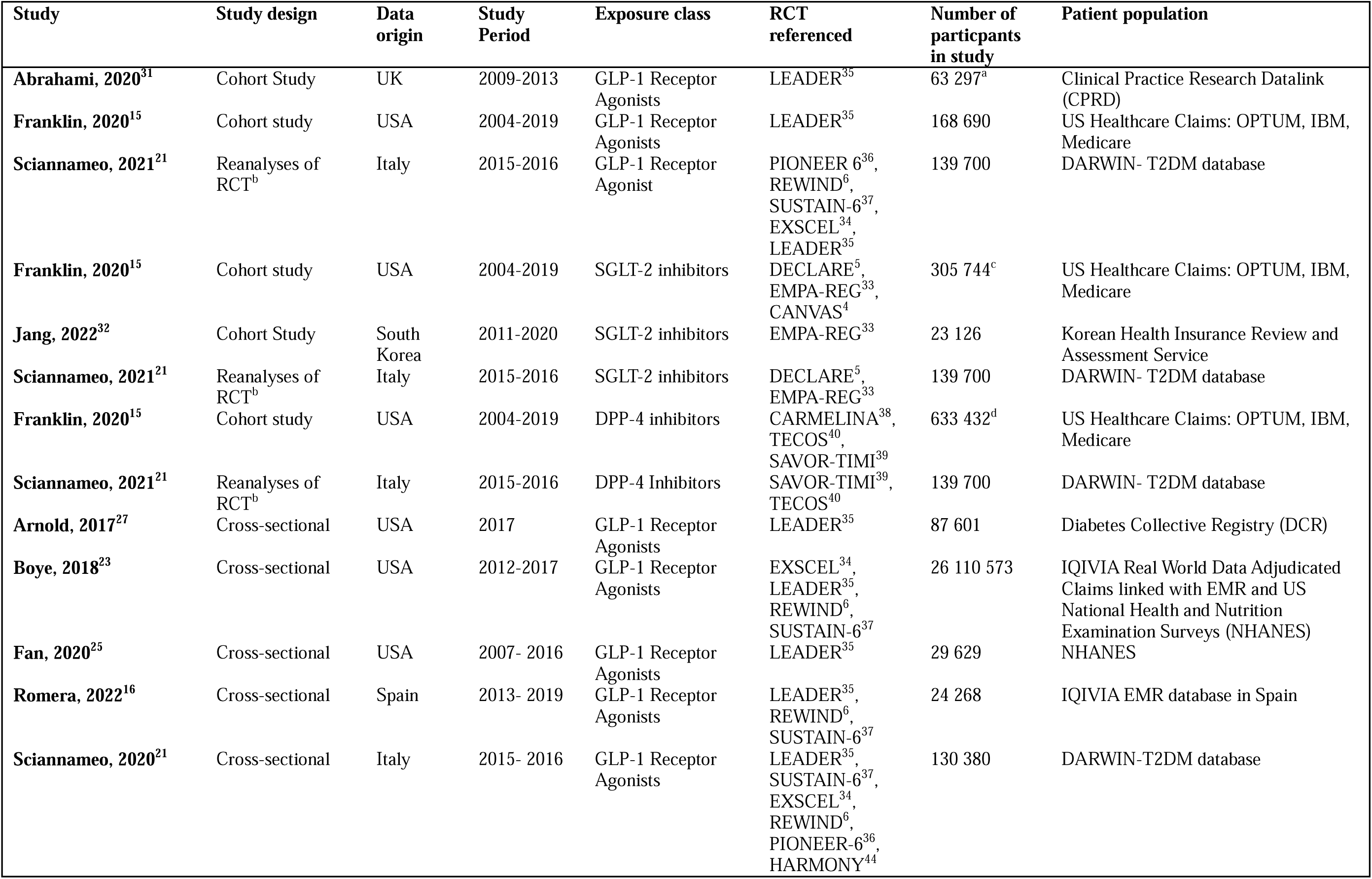
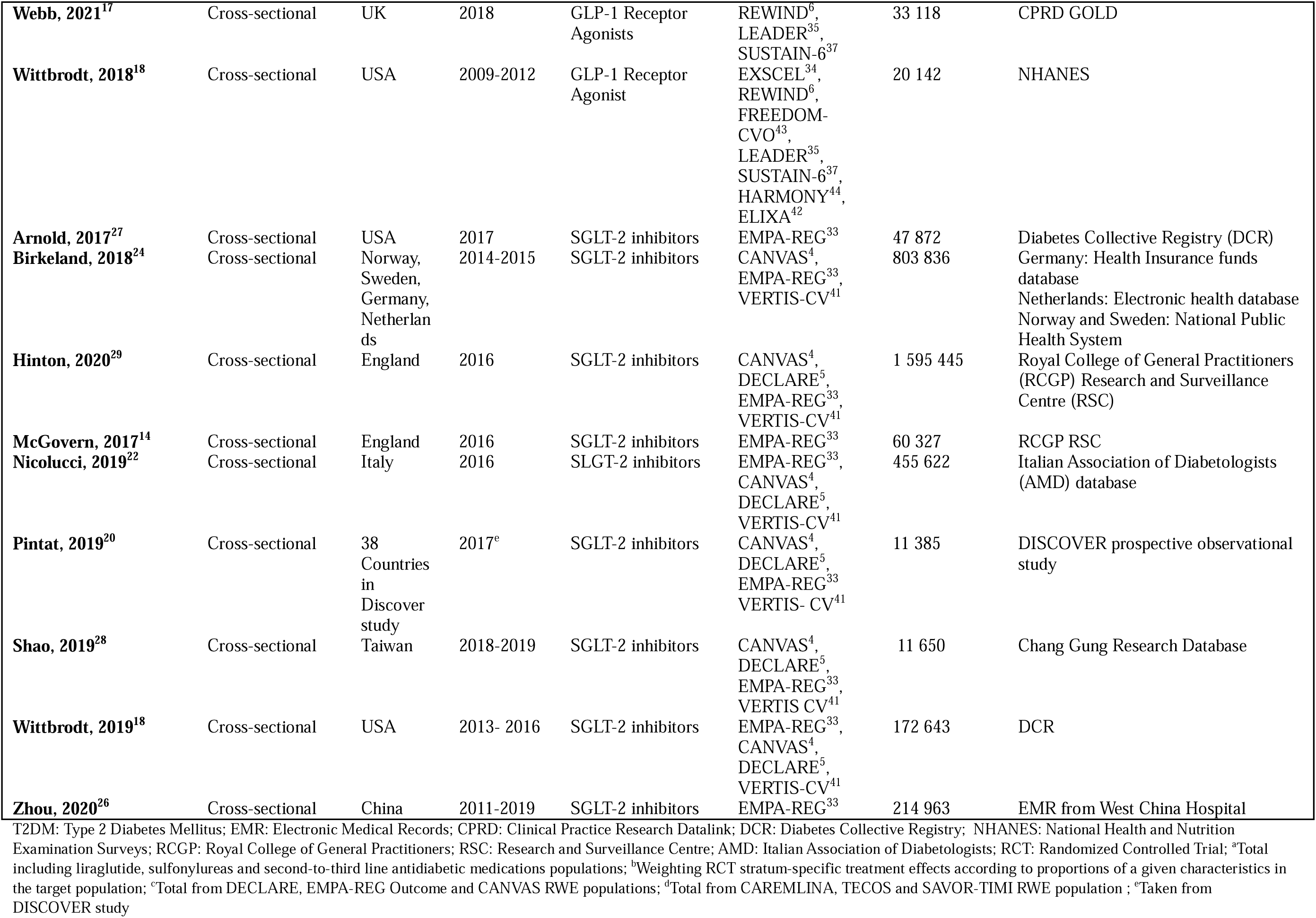
Summary of study characteristics for systematic review of observational studies replicating cardiovascular outcome trials among patients with type 2 diabetes.

| Study | Study design | Data origin | Study Period | Exposure class | RCT referenced | Number of participants in study | Patient population |
| --- | --- | --- | --- | --- | --- | --- | --- |
| **Abrahami, 2020<sup>31</sup>** | Cohort Study | UK | 2009-2013 | GLP-1 Receptor Agonists | LEADER<sup>35</sup> | 63 297<sup>a</sup> | Clinical Practice Research Datalink (CPRD) |
| **Franklin, 2020<sup>15</sup>** | Cohort study | USA | 2004-2019 | GLP-1 Receptor Agonists | LEADER<sup>35</sup> | 168 690 | US Healthcare Claims: OPTUM, IBM, Medicare |
| **Sciannameo, 2021<sup>21</sup>** | Reanalyses of RCT<sup>b</sup> | Italy | 2015-2016 | GLP-1 Receptor Agonist | PIONEER-6<sup>36</sup>,<br>REWIND<sup>6</sup>,<br>SUSTAIN-6<sup>37</sup>,<br>EXSCEL<sup>34</sup>,<br>LEADER<sup>35</sup> | 139 700 | DARWIN- T2DM database |
| **Franklin, 2020<sup>15</sup>** | Cohort study | USA | 2004-2019 | SGLT-2 inhibitors | DECLARE<sup>5</sup>,<br>EMPA-REG<sup>33</sup>,<br>CANVAS<sup>4</sup> | 305 744<sup>c</sup> | US Healthcare Claims: OPTUM, IBM, Medicare |
| **Jang, 2022<sup>32</sup>** | Cohort Study | South Korea | 2011-2020 | SGLT-2 inhibitors | EMPA-REG<sup>33</sup> | 23 126 | Korean Health Insurance Review and Assessment Service |
| **Sciannameo, 2021<sup>21</sup>** | Reanalyses of RCT<sup>b</sup> | Italy | 2015-2016 | SGLT-2 inhibitors | DECLARE<sup>5</sup>,<br>EMPA-REG<sup>33</sup> | 139 700 | DARWIN- T2DM database |
| **Franklin, 2020<sup>15</sup>** | Cohort study | USA | 2004-2019 | DPP-4 inhibitors | CARMELINA<sup>38</sup>,<br>TECOS<sup>40</sup>,<br>SAVOR-TIMI<sup>39</sup> | 633 432<sup>d</sup> | US Healthcare Claims: OPTUM, IBM, Medicare |
| **Sciannameo, 2021<sup>21</sup>** | Reanalyses of RCT<sup>b</sup> | Italy | 2015-2016 | DPP-4 Inhibitors | SAVOR-TIMI<sup>39</sup>,<br>TECOS<sup>40</sup> | 139 700 | DARWIN- T2DM database |
| **Arnold, 2017<sup>27</sup>** | Cross-sectional | USA | 2017 | GLP-1 Receptor Agonists | LEADER<sup>35</sup> | 87 601 | Diabetes Collective Registry (DCR) |
| **Boye, 2018<sup>23</sup>** | Cross-sectional | USA | 2012-2017 | GLP-1 Receptor Agonists | EXSCEL<sup>34</sup>,<br>LEADER<sup>35</sup>,<br>REWIND<sup>6</sup>,<br>SUSTAIN-6<sup>37</sup> | 26 110 573 | IQIVIA Real World Data Adjudicated Claims linked with EMR and US National Health and Nutrition Examination Surveys (NHANES) |
| **Fan, 2020<sup>25</sup>** | Cross-sectional | USA | 2007- 2016 | GLP-1 Receptor Agonists | LEADER<sup>35</sup> | 29 629 | NHANES |
| **Romera, 2022<sup>16</sup>** | Cross-sectional | Spain | 2013- 2019 | GLP-1 Receptor Agonists | LEADER<sup>35</sup>,<br>REWIND<sup>6</sup>,<br>SUSTAIN-6<sup>37</sup> | 24 268 | IQIVIA EMR database in Spain |
| **Sciannameo, 2020<sup>21</sup>** | Cross-sectional | Italy | 2015- 2016 | GLP-1 Receptor Agonists | LEADER<sup>35</sup>,<br>SUSTAIN-6<sup>37</sup>,<br>EXSCEL<sup>34</sup>,<br>REWIND<sup>6</sup>,<br>PIONEER-6<sup>36</sup>,<br>HARMONY<sup>44</sup> | 130 380 | DARWIN-T2DM database |
| **Webb, 2021<sup>17</sup>** | Cross-sectional | UK | 2018 | GLP-1 Receptor Agonists | REWIND<sup>6</sup>, LEADER<sup>35</sup>, SUSTAIN-6<sup>37</sup> | 33 118 | CPRD GOLD |
| **Wittbrodt, 2018<sup>18</sup>** | Cross-sectional | USA | 2009-2012 | GLP-1 Receptor Agonist | EXSCEL<sup>34</sup>, REWIND<sup>6</sup>, FREEDOM-CVO<sup>43</sup>, LEADER<sup>35</sup>, SUSTAIN-6<sup>37</sup>, HARMONY<sup>44</sup>, ELIXA<sup>42</sup> | 20 142 | NHANES |
| **Arnold, 2017<sup>27</sup>** | Cross-sectional | USA | 2017 | SGLT-2 inhibitors | EMPA-REG<sup>33</sup> | 47 872 | Diabetes Collective Registry (DCR) |
| **Birkeland, 2018<sup>24</sup>** | Cross-sectional | Norway, Sweden, Germany, Netherlands | 2014-2015 | SGLT-2 inhibitors | CANVAS<sup>4</sup>, EMPA-REG<sup>33</sup>, VERTIS-CV<sup>41</sup> | 803 836 | Germany: Health Insurance funds database<br>Netherlands: Electronic health database<br>Norway and Sweden: National Public Health System |
| **Hinton, 2020<sup>29</sup>** | Cross-sectional | England | 2016 | SGLT-2 inhibitors | CANVAS<sup>4</sup>, DECLARE<sup>5</sup>, EMPA-REG<sup>33</sup>, VERTIS-CV<sup>41</sup> | 1 595 445 | Royal College of General Practitioners (RCGP) Research and Surveillance Centre (RSC) |
| **McGovern, 2017<sup>14</sup>** | Cross-sectional | England | 2016 | SGLT-2 inhibitors | EMPA-REG<sup>33</sup> | 60 327 | RCGP RSC |
| **Nicolucci, 2019<sup>22</sup>** | Cross-sectional | Italy | 2016 | SGLT-2 inhibitors | EMPA-REG<sup>33</sup>, CANVAS<sup>4</sup>, DECLARE<sup>5</sup>, VERTIS-CV<sup>41</sup> | 455 622 | Italian Association of Diabetologists (AMD) database |
| **Pintat, 2019<sup>20</sup>** | Cross-sectional | 38 Countries in Discover study | 2017<sup>c</sup> | SGLT-2 inhibitors | CANVAS<sup>4</sup>, DECLARE<sup>5</sup>, EMPA-REG<sup>33</sup>, VERTIS-CV<sup>41</sup> | 11 385 | DISCOVER prospective observational study |
| **Shao, 2019<sup>28</sup>** | Cross-sectional | Taiwan | 2018-2019 | SGLT-2 inhibitors | CANVAS<sup>4</sup>, DECLARE<sup>5</sup>, EMPA-REG<sup>33</sup>, VERTIS CV<sup>41</sup> | 11 650 | Chang Gung Research Database |
| **Wittbrodt, 2019<sup>18</sup>** | Cross-sectional | USA | 2013- 2016 | SGLT-2 inhibitors | EMPA-REG<sup>33</sup>, CANVAS<sup>4</sup>, DECLARE<sup>5</sup>, VERTIS-CV<sup>41</sup> | 172 643 | DCR |
| **Zhou, 2020<sup>26</sup>** | Cross-sectional | China | 2011-2019 | SGLT-2 inhibitors | EMPA-REG<sup>33</sup> | 214 963 | EMR from West China Hospital |

T2DM: Type 2 Diabetes Mellitus; EMR: Electronic Medical Records; CPRD: Clinical Practice Research Datalink; DCR: Diabetes Collective Registry; NHANES: National Health and Nutrition Examination Surveys; RCGP: Royal College of General Practitioners; RSC: Research and Surveillance Centre; AMD: Italian Association of Diabetologists; RCT: Randomized Controlled Trial; <sup>a</sup>Total including liraglutide, sulfonylureas and second-to-third line antidiabetic medications populations; <sup>b</sup>Weighting RCT stratum-specific treatment effects according to proportions of a given characteristics in the target population; <sup>c</sup>Total from DECLARE, EMPA-REG Outcome and CANVAS RWE populations; <sup>d</sup>Total from CAREMLINA, TECOS and SAVOR-TIMI RWE population ; <sup>e</sup>Taken from DISCOVER study

### MACE Outcomes in Emulated RCTs

Four studies emulated RCTs using RWD (**Table 2**). Sciannameo et al.^21^ used odds weighting based on RCT subgroup-specific HRs to transport results onto their RWD population; consequently, there was no real-world exposure or comparator for these analyses. As the exposures and outcomes in these analyses were directly from the relevant RCTs, we did not calculate the agreement statistics for this study. For the other three studies, exposure definitions of intention-to-treat (ITT) or on-treatment were used with an active comparator. These studies also imitated the inclusion criteria and exclusion criteria of the emulated RCTs. They also ensured follow-up was the same duration as the RCT and used propensity score matching to reduce confounding. MACE was defined as a composite endpoint of cardiovascular or all-cause death, stroke, and MI; Franklin et al. emulated TECOS by including angina in their MACE definition^11^.

**Table 2.** Summary of MACE outcomes for RWD and RCT studies grouped by exposure class. Abbreviations: HR: Hazard Ratio; CI: Confidence Interval; ITT: intention-to-treat, DPP-4i: Dipeptidyl peptidase 4 inhibitors; SU: sulfonylureas; 2^nd^ – gen SU: Second generation sulfonylureas; RCT: Randomized Controlled Trial; ^a^SA: Full statistical significance agreement (Y/N/P) – determines if the RWD and RCT have estimates and CIs on the same side of the null; P: partial significance agreement-met the prespecified noninferiority criteria even though the database study may have indicated superiority, EA: Estimate agreement (Y/N) – determines if the effect estimate of the RWD falls within the 95% confidence interval of the RCT effect estimate. SD: Standardized difference agreement is defined by standardized differences |Z| < 1.96 (Y = yes, N = no). ^b^ Weighting RCT stratum-specific treatment effects according to proportions of a given characteristics in the target population; ^c^second to third line antidiabetic drugs; ^d^ Calculated by hand; ^e^MACE including angina;

| Study | RCT | Exposure definition | Exposure | Comparator | Exposure N | Comparat or N | HR | 95% CI | RCT HR | 95% CI | Observed Agreement<sup>a</sup> |  |  |
| --- | --- | --- | --- | --- | --- | --- | --- | --- | --- | --- | --- | --- | --- |
|  |  |  |  |  |  |  |  |  |  |  | SA | EA | SD |
| **<u>SGLT-2 inhibitors</u>** |  |  |  |  |  |  |  |  |  |  |  |  |  |
| Jang, 2022 | EMPA-REG<sup>33</sup> | ITT | Empagliflozin | Sitagliptin | 11 563 | 11 563 | 0.87 | 0.79, 0.96 | 0.86 | 0.74, 0.99 | Y | Y | Y |
| Franklin, 2020 | EMPA-REG<sup>33</sup> | ITT | Empagliflozin | DPP-4i | 4 687 | 2 333 | 0.83 | 0.73, 0.94 | 0.86 | 0.74, 0.99 | Y | Y | Y |
| Franklin, 2020 | CANVAS<sup>4</sup> | On treatment | Canagliflozin | DPP-4i | 5 795 | 4 347 | 0.77 | 0.70, 0.85 | 0.86 | 0.75, 0.97 | Y | Y | Y |
| Sciannameo, 2021<sup>b</sup> | DECLARE<sup>5</sup> | N/A | N/A | N/A | N/A | N/A | 0.94 | 0.84, 1.04 | 0.93 | 0.84, 1.03 | - | - | - |
| Sciannameo, 2021<sup>b</sup> | EMPA-REG<sup>33</sup> | N/A | N/A | N/A | N/A | N/A | 0.88 | 0.74, 1.03 | 0.86 | 0.74, 0.99 | - | - | - |
| **<u>GLP-1 Receptor Agonists</u>** |  |  |  |  |  |  |  |  |  |  |  |  |  |
| Abrahami, 2020 | LEADER<sup>35</sup> | ITT | Liraglutide | SU | 1 868 | 25 895 | 1.03 | 0.82, 1.30 | 0.87 | 0.78, 0.97 | N | N | Y<sup>d</sup> |
| Abrahami, 2020 | LEADER<sup>35</sup> | ITT | Liraglutide | 2<sup>nd</sup> – 3<sup>rd</sup> line<sup>c</sup> | 1 864 | 32 899 | 0.97 | 0.78, 1.22 | 0.87 | 0.78, 0.97 | N | Y | Y<sup>d</sup> |
| Franklin, 2020 | LEADER<sup>35</sup> | On treatment | Liraglutide | DPP-4i | 4 668 | 4 672 | 0.82 | 0.76, 0.87 | 0.87 | 0.78, 0.97 | Y | Y | Y |
| Sciannameo, 2021<sup>b</sup> | EXSCEL<sup>34</sup> | N/A | N/A | N/A | N/A | N/A | 0.92 | 0.82, 1.02 | 0.91 | 0.83, 1.00 | - | - | - |
| Sciannameo, 2021<sup>b</sup> | LEADER<sup>35</sup> | N/A | N/A | N/A | N/A | N/A | 0.88 | 0.77, 0.99 | 0.87 | 0.78, 0.97 | - | - | - |
| Sciannameo, 2021<sup>b</sup> | PIONEER-6<sup>36</sup> | N/A | N/A | N/A | N/A | N/A | 0.76 | 0.41, 1.10 | 0.79 | 0.57, 1.11 | - | - | - |
| Sciannameo, 2021<sup>b</sup> | REWIND<sup>6</sup> | N/A | N/A | N/A | N/A | N/A | 0.87 | 0.76, 0.98 | 0.88 | 0.79, 0.99 | - | - | - |
| Sciannameo, 2021<sup>b</sup> | SUSTAIN-6<sup>37</sup> | N/A | N/A | N/A | N/A | N/A | 0.73 | 0.47, 0.99 | 0.74 | 0.58, 0.95 | - | - | - |
| **<u>DPP-4 Inhibitors</u>** |  |  |  |  |  |  |  |  |  |  |  |  |  |
| Franklin, 2020 | CARMELIN<sup>38</sup> | On treatment | Linagliptin | 2<sup>nd</sup> – gen SU | 3 494 | 3 485 | 0.90 | 0.84, 0.96 | 1.02 | 0.89, 1.17 | P | Y | Y |
| Franklin, 2020 | SAVOR-TIMI<sup>39</sup> | On treatment | Saxagliptin | 2<sup>nd</sup> – gen SU | 8 280 | 8 212 | 0.81 | 0.76, 0.86 | 1.00 | 0.89, 1.12 | P | N | Y |
| Franklin, 2020<sup>e</sup> | TECOS<sup>40</sup> | On treatment | Sitagliptin | 2<sup>nd</sup> – gen SU | 7 257 | 7 266 | 0.89 | 0.86, 0.91 | 0.98 | 0.88, 1.09 | P | Y | Y |
| Sciannameo, 2021<sup>b</sup> | SAVOR-TIMI<sup>39</sup> | N/A | N/A | N/A | N/A | N/A | 0.99 | 0.87, 1.10 | 1.00 | 0.89, 1.12 | - | - | - |
| Sciannameo, 2021<sup>b</sup> | TECOS<sup>40</sup> | N/A | N/A | N/A | N/A | N/A | 0.97 | 0.87, 1.06 | 0.98 | 0.88, 1.09 | - | - | - |

For studies that investigated SGLT-2 inhibitors, there were three RCTs examined: EMPA-REG Outcome^33^, CANVAS^4^ and DECLARE^5^. Treatment matched the SGLT-2 inhibitors evaluated in the RCTs. However, while the RCTs used a placebo comparator, active comparators (DPP-4 inhibitors) were used in the RWD studies. There was strong agreement between RWD and RCTs, with the three studies achieving full statistical significance agreement and estimate agreement, with HRs from the RWD in the same direction as those from the RCTs and within the RCT’s 95% CIs.

There were three analyses of GLP-1 receptor agonists, which examined six RCTs (EXSCEL^34^, LEADER^35^, PIONEER-6^36^, REWIND^6^, SUSTAIN-6^37^). The RWD studies used ITT and on-treatment exposure definitions. For the emulation of the LEADER trial, liraglutide was compared with sulfonylureas, second-to-third-line antidiabetic drugs, and DPP-4 inhibitors. Across the three different analyses, all showed partial statistical significance agreement. All studies except Abrahami et al. evaluating liraglutide compared to sulfonylureas demonstrated estimate agreement^31^.

There were five analyses of DPP-4 inhibitors, which examined three RCTs (CARMELINA^38^, SAVOR-TIMI-53^39^, and TECOS^40^). RCT odds weighting was used in two of the five analyses. In the other three analyses, an on-treatment exposure definition was used, and second-generation sulfonylureas were used as the comparator. All analyses were able to replicate the non-inferiority findings of the RCTs, and all but one demonstrated estimate agreement.

### Secondary Clinical Outcomes in Emulated RCTs

**Table 3** summarizes the results for secondary clinical outcomes in the RWD studies and the emulated RCTs. Jang et al, which emulated the EMPA-REG Outcome trial by comparing empagliflozin with sitagliptin, examined select individual components of MACE (MI and stroke), all-cause mortality, and HHF^32^. All-cause mortality, MI, and HHF had either partial or full statistical significance agreement and estimate agreement. Empagliflozin was found to be non-inferior for stroke in RWD but not the RCT; thus, statistical significance agreement was not met for this outcome. However, there was estimate agreement^32^. Sciannameo et al. and Franklin et al. examined HHF and/or cardiovascular death when replicating the DECLARE trial^15,21^. Franklin et al. found both regulatory agreement for superiority and estimate agreement.

**Table 3.** Summary of secondary outcomes of all-cause mortality, MI, Stroke, HHF and CV death for RWD and RCT studies for SGLT-2 Inhibitors. Abbreviations: HR: Hazard Ratio; CI: Confidence Interval; ITT: intention-to-treat; HHF: hospitalization due to heart failure; CV: cardiovascular; RCT: Randomized Controlled Trial; DPP-4i: Dipeptidyl peptidase 4 inhibitors ^a^ SA: Full statistical significance agreement (Y/N/P) – determines if the RWD and RCT have estimates and CIs on the same side of the null; P: partial significance agreement-met the prespecified noninferiority criteria even though the database study may have indicated superiority, EA: Estimate agreement (Y/N) – determines if the effect estimate of the RWD falls within the 95% confidence interval of the RCT effect estimate. SD: Standardized difference agreement is defined by standardized differences |Z| < 1.96 (Y = yes, N = no). ^b^ Calculated by hand; ^c^Weighting RCT stratum-specific treatment effects according to proportions of a given characteristics in the target population.

| Study | RCT | Exposure definition | Exposure | Comparator | Exposure N | Comparator N | Outcome | HR | 95% CI | RCT HR | RCT 95% CI | Observed Agreement <sup>a</sup> |  |  |
| --- | --- | --- | --- | --- | --- | --- | --- | --- | --- | --- | --- | --- | --- | --- |
|  |  |  |  |  |  |  |  |  |  |  |  | SA | EA | SD |
| **Jang, 2022** | EMPA-REG<sup>33</sup> | ITT | Empagliflozin | Sitagliptin | 11 563 | 11 563 | All cause death | 0.78 | 0.67, 0.91 | 0.68 | 0.57, 0.82 | Y | Y | Y |
| **Jang, 2022** | EMPA-REG<sup>33</sup> | ITT | Empagliflozin | Sitagliptin | 11 563 | 11 563 | MI | 0.91 | 0.76, 1.08 | 0.87 | 0.70, 1.09 | Y | Y | Y |
| **Jang, 2022** | EMPA-REG<sup>33</sup> | ITT | Empagliflozin | Sitagliptin | 11 563 | 11 563 | Stroke | 0.89 | 0.75, 1.05 | 1.18 | 0.89, 1.56 | N | Y | Y |
| **Jang, 2022** | EMPA-REG<sup>33</sup> | ITT | Empagliflozin | Sitagliptin | 11 563 | 11 563 | HHF | 0.85 | 0.75, 0.95 | 0.65 | 0.50, 0.85 | Y | Y | Y |
| **Franklin, 2020** | DECLARE<sup>5</sup> | ITT | Dapagliflozin | DPP-4i | 8 582 | 8 578 | HHF and CV death | 0.70 | 0.59, 0.82 | 0.83 | 0.73, 0.95 | Y | Y | Y<sup>b</sup> |
| **Sciannameo, 2021<sup>c</sup>** | DECLARE<sup>5</sup> | N/A | N/A | N/A | N/A | N/A | HHF and/or CV death | 0.86 | 0.73, 0.99 | 0.83 | 0.73, 0.95 | - | - | - |

### Eligibility

**Table 4** summarizes the findings when examining the proportion of RWD population eligible (including cross-sectional studies) for CVOTs. In total, there were 59 populations across CVOTs evaluating SGLT-2 inhibitors and GLP-1 receptor agonists. Populations varied in composition, with some from the general population, others only from select populations (inpatient vs outpatient), and others restricted to users of the drug class of interest.

**Table 4.**
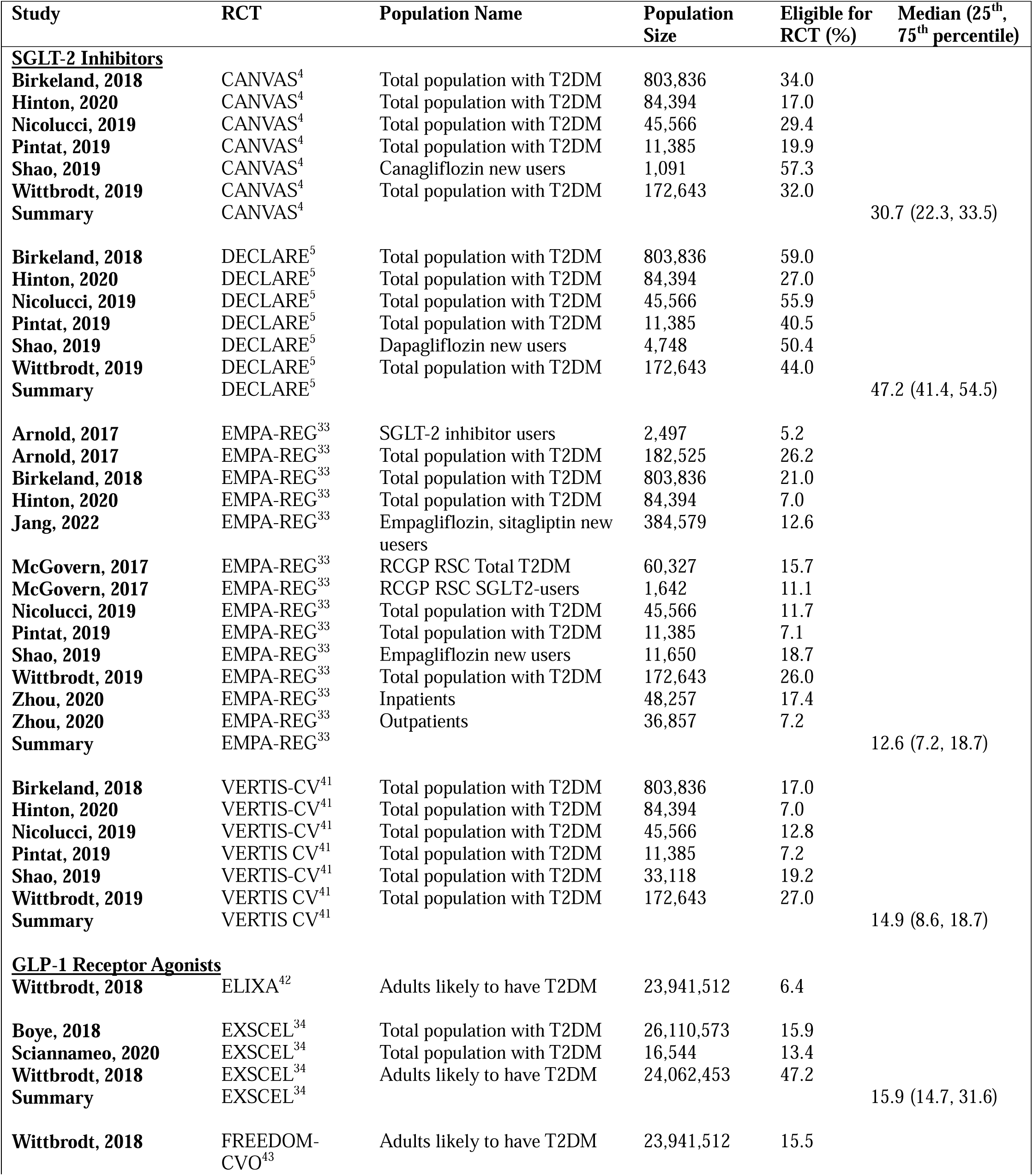
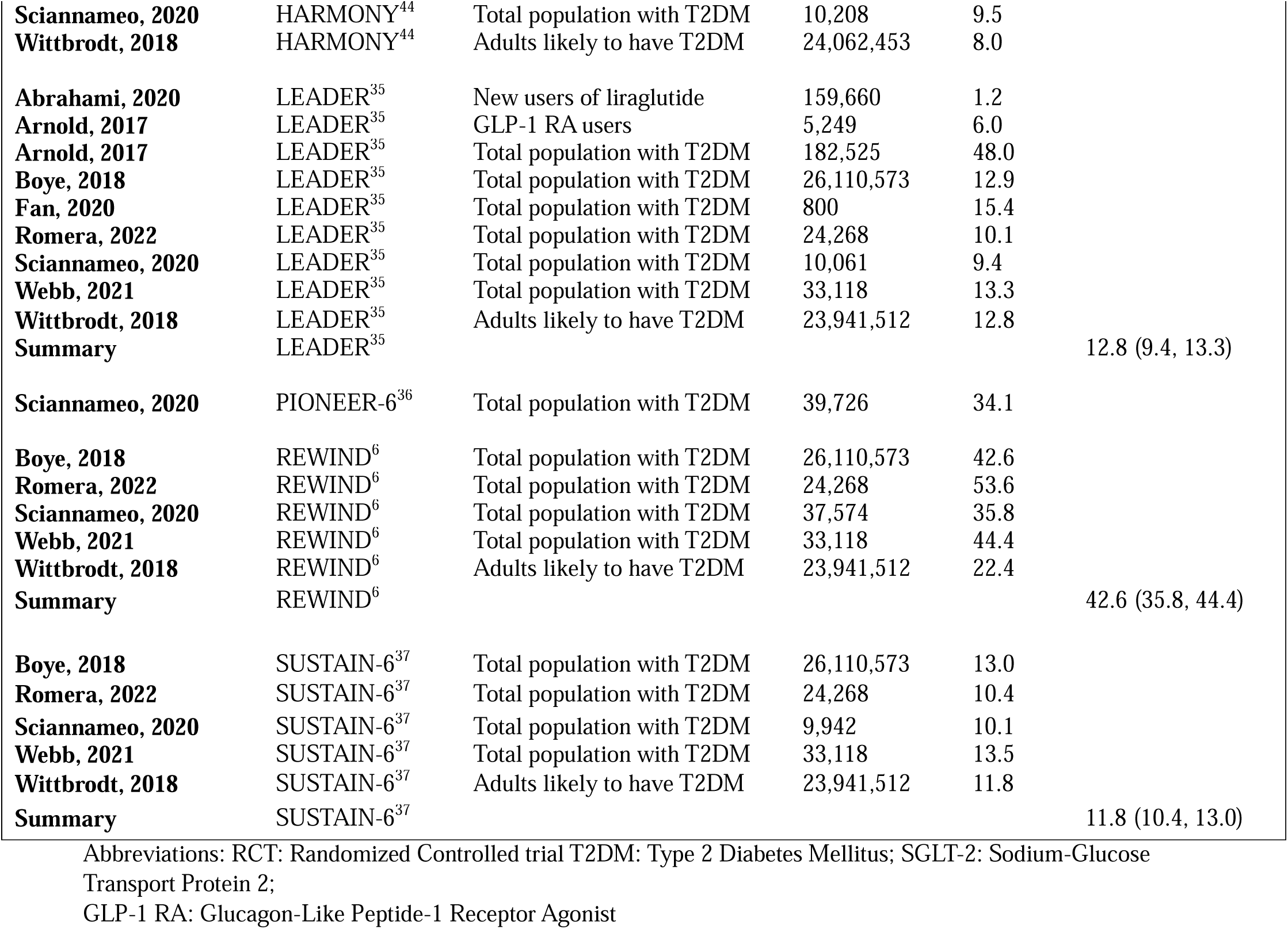
Summary of proportion of real-world patients eligible for cardiovascular outcome trials examining newer antidiabetic drugs.

For SGLT-2 inhibitors, RWD populations were compared to a total of four RCTs (CANVAS^4^, DECLARE^5^, EMPA-REG Outcome,^33^ and VERTIS-CV^41^). The median percentage eligible from the trials ranged between 12.6% to 30.7%. Of note, the Jang et al. study that demonstrated full statistical, estimate, and standardized difference agreement with the EMPA-REG Outcome trial had an 12.6% eligible for the RCT from a population of patients with T2DM who were new-users of empagliflozin or sitagliptin. For GLP-1 receptor agonists, RWD populations were compared to eight different RCTs (ELIXA^42^, EXSCEL^34^, FREEDOM-CVO^43^, HARMONY^44^, LEADER^35^, PIONEER-6^36^, REWIND^6^, SUSTAIN-6^37^). Median percentage eligible from the trials ranged from 11.8% to 42.6%. No included studies evaluated trial eligibility for DPP-4 inhibitors.

We conducted post-hoc analyses for patient characteristics which can be found in **Figures S1-S7**. There were no association between age or sex in RWD populations and percentage of individuals who were eligible for the RCTs. Moreover, there were substantial differences in patient characteristics such as age, sex, use of antidiabetic medication between the RWD populations compared to the populations examined in the respective RCTs.

## DISCUSSION

Our study was designed to synthesize observational studies that emulated CVOTs in patients with T2DM and to summarize cross-sectional studies assessing the proportion of real-world populations eligible for previous CVOTs. We found strong agreement between results from RWD and RCTs for SGLT-2 inhibitors, GLP-1 receptor agonists, and DPP-4 inhibitors. Individual components of MACE, HHF, and cardiovascular death also showed consistency between the RWD studies^15,21,32^ and the SGLT-2 inhibitor CVOTs, EMPA-REG Outcome^33^ and DECLARE^5^. There was considerable heterogeneity in the percentage of individuals who would have been eligible for the CVOTs of SGLT-2 inhibitors and GLP-1 receptor agonists. Patient characteristics also varied between RWD and RCTs, with no association between patient characteristics and real-world proportion eligible for RCTs.

Our systematic review highlighted the heterogeneity of methods used to emulate RCTs using RWD. As this is an emerging area of research, the studies included in our study had different exposure definitions, active comparators, and methods to reduce confounding. The lack of agreement between Abrahami et al’s study and the LEADER trial highlights how differences in comparators from sulfonylureas to broader 2^nd^ or 3^rd^ line antidiabetic drugs influenced the direction of the effect estimate^31^. In addition, some studies utilized an ITT approach where exposure was defined at cohort entry until end of follow-up while others used an on-treatment approach which censored for treatment discontinuation or switching. As these changes may impact the research question and effect estimate, more research is needed to better understand the strengths and limitations of using different methods for replicating RCTs with RWD.

Our study has shown that studies emulating RCTs using RWD to estimate the effects of newer antidiabetic medications on MACE generally demonstrated agreement. However, there was some heterogeneity in our results, which may be due to the types of methods used for replication such as choice of exposure definition (ITT vs on-treatment), choice of active comparator, and methods to adjust for confounding. In addition, there were some differences between RCTs and RWD which may be due to inherent differences between the two designs, such as the source population, exposure (active comparator vs placebo), follow-up duration, and how events are captured and measured. The results may have also differed due to chance. Overall, our systematic review demonstrated that similar conclusions can be obtained when using RWD to emulate RCTs when efforts are made in study design to closely mimic RCTs. While RCTs are the gold-standard for the generation of evidence, regulatory bodies in countries/regions such as Canada, USA and the European Union are increasingly evaluating the use of RWD in decision-making^2,45,46^. Our results provide reassurance that with the proper methods, RWD can similarly generate quality evidence.

The proportion of the real-world population that would have been eligible for the RCTs varied across studies but was almost always less than 50%. The lack of generalizability of these RCT populations is driven in part by their design. These trials predominantly included older individuals who possessed one or more cardiovascular disease (CVD) risk factors and generally had longer durations of disease. These factors were necessary for event driven CVOTs to ensure that that a sufficient number of events could be accrued in a short time frame^3^. This requirement explains in part the observed heterogeneity (and large standardized mean differences for patient characteristics) between the RWD and the RCT populations. For example, the RWD populations consistently had lower percentages of established CVD. Given these differences, it is important to utilize studies from routine practice settings to complement findings from highly controlled RCTs to account for differences in patients and practice for clinical decision making^3^. There are opportunities to use RWD for Phase IV trials and pragmatic trials as highlighted by the FDA^2^.

Previous reviews in the area of RCT emulation using RWD examined the reporting of RWD studies that aimed to replicate RCTs^47,48^. They found inconsistencies in the reporting of key elements in these studies. For example, in a systematic review of 200 studies aiming to emulate RCTs using observational data, Hansford et al. reported that 43% did not describe all key elements of how the target trial was emulated^47^. In addition, only 37% of studies reported potential unmeasured confounders. A scoping review of 96 studies aiming to emulate a target trial across medical fields also identified unmeasured confounding as the most commonly stated limitation^48^. We found that the cohort studies included in our systematic review all reported the key elements of eligibility criteria, treatment strategy, assignment procedures, outcome, follow-up, causal contrast, analyses plan and specification of time-zero. However, they did not comment on potential unmeasured confounders. Our study builds upon these previous studies by examining agreement statistics between RWD and RCTs, the proportion of RWD patients who would have been eligible for RCTs, and differences in patient characteristics. The recent development of guidelines for the reporting of target trial emulation may improve reporting in this area^49^.

Our study has many strengths. It included a comprehensive, systematic search that was constructed with the assistance of an experienced librarian. To our knowledge, this is the first systematic review to examine the emulation of CVOTs for newer antidiabetic drugs using RWD. In addition, we assessed the agreement between the RWD studies and RCTs based on regulatory standards. Furthermore, our study is the first to synthesize the generalizability of CVOTs in the real-world by examining the proportion eligible of real-world populations for these CVOTs. We also compared patient characteristics between the CVOTs and RWD to better understand differences in study populations.

Our study has several potential limitations. First, there was a limited number of studies emulating RCTs using RWD among patients with T2DM. These studies also had important heterogeneity in study design, drug class, and analytical approach. Consequently, we conducted a systematic review without formal meta-analysis. Second, as an emerging area of research, there are currently no established guidelines on how to assess the quality of these studies for external validity. While we used the DUPLICATE Team’s agreement statistics^12,15^ to assess the estimates achieved from the emulation in comparison to the RCT that was being emulated, these statistics have their own limitations. For example, it is difficult to replicate studies that have shown a null effect, as observational studies often have more precision. This issue is well illustrated by our results for DPP-4 inhibitors where RWD studies achieved superiority while the RCTs reported a null effect^15,21^. Moreover, the estimate agreement is contingent on the precision of the RCT. If the RCT has a wider CI, there is a higher probability that the RWD estimate will fall within this interval. Third, to our knowledge, there are no guidelines or standards on reporting or assessing external validity of RCT. Finally, we chose to include cross-sectional studies for our secondary objectives examining the proportion eligible of real-world populations for RCTs and their characteristics, however, these studies had considerable missing data and are at an inherently higher risk of bias.

## AUTHORS’ CONTRIBUTIONS

WW led the protocol development, conducted the analysis, and drafted the manuscript. WW and WT extracted data. All authors contributed to protocol development, were involved in data interpretation, critically reviewed the manuscript for important intellectual content, and approved the final manuscript. KBF supervised the study and is the guarantor.

## Data Availability

All data produced in the present study are available upon reasonable request to the authors

## Supplementary Materials Titles

**Table S1** PRISMA Checklist for systematic review of observational studies assessing generalizability of CVOTS

**Table S2** SWiM Reporting Checklist for systematic review of observational studies assessing generalizability of CVOTS

**Table S3** Search strategy for systematic review of observational studies assessing generalizability of CVOTS using OVID MEDLINE database

**Table S4** Search strategy for systematic review of observational studies assessing generalizability of CVOTS using OVID EMBASE + EMBASE CLASSIC database

**Table S5** Search strategy for systematic review of observational studies assessing generalizability of CVOTS using Cochrane CENTRAL database

**Table S6** A list of completed cardiovascular outcome trials and their drug class and molecule

**Table S7** Standardized differences computed for observational study compared to RCT hazard ratios

**Figure S1** Distribution of percent eligible for inclusion in CVOT of SGLT-2 inhibitors in real-world populations by mean age (i) and by sex (ii)

**Figure S2** Distribution of percent eligible for inclusion in CVOT of GLP-1 Receptor Agonists in real-world populations by mean age (i) and by sex (ii)

**Figure S3** Standardized Mean Differences of patient characteristics between GLP-1 Receptor Agonist CVOTs and the study population from the RWD study from Sciannameo, 2020

**Figure S4** Select standardized mean difference of patient characteristics for CVOT and RWD for the DECLARE trial

**Figure S5** Select standardized mean difference of patient characteristics for CVOT and RWD for CANVAS trial

