## Supplementary Materials for "Replicating cardiovascular outcome trials of medications used to treat type 2 diabetes using real-world data: A systematic review of observational studies"

**Table S1** PRISMA Checklist for systematic review of observational studies assessing generalizability of CVOTS

| **Section and Topic** | **Item #** | **Checklist item** | **Location where item is reported (Page)** |
| --- | --- | --- | --- |
| **TITLE** | | |  |
| Title | 1 | Identify the report as a systematic review. | 1 |
| **ABSTRACT** | | |  |
| Abstract | 2 | See the PRISMA 2020 for Abstracts checklist. | 2 |
| **INTRODUCTION** | | |  |
| Rationale | 3 | Describe the rationale for the review in the context of existing knowledge. | 3, 4 |
| Objectives | 4 | Provide an explicit statement of the objective(s) or question(s) the review addresses. | 4 |
| **METHODS** | | |  |
| Eligibility criteria | 5 | Specify the inclusion and exclusion criteria for the review and how studies were grouped for the syntheses. | 5, 7 |
| Information sources | 6 | Specify all databases, registers, websites, organisations, reference lists and other sources searched or consulted to identify studies. Specify the date when each source was last searched or consulted. | 4 |
| Search strategy | 7 | Present the full search strategies for all databases, registers and websites, including any filters and limits used. | Supplementary Material |
| Selection process | 8 | Specify the methods used to decide whether a study met the inclusion criteria of the review, including how many reviewers screened each record and each report retrieved, whether they worked independently, and if applicable, details of automation tools used in the process. | 4,5 |
| Data collection process | 9 | Specify the methods used to collect data from reports, including how many reviewers collected data from each report, whether they worked independently, any processes for obtaining or confirming data from study investigators, and if applicable, details of automation tools used in the process. | 6 |
| Data items | 10a | List and define all outcomes for which data were sought. Specify whether all results that were compatible with each outcome domain in each study were sought (e.g. for all measures, time points, analyses), and if not, the methods used to decide which results to collect. | 5 |
|  | 10b | List and define all other variables for which data were sought (e.g. participant and intervention characteristics, funding sources). Describe any assumptions made about any missing or unclear information. | 5 |
| Study risk of bias assessment | 11 | Specify the methods used to assess risk of bias in the included studies, including details of the tool(s) used, how many reviewers assessed each study and whether they worked independently, and if applicable, details of automation tools used in the process. | N/A |
| Effect measures | 12 | Specify for each outcome the effect measure(s) (e.g. risk ratio, mean difference) used in the synthesis or presentation of results. | 7 |
| Synthesis methods | 13a | Describe the processes used to decide which studies were eligible for each synthesis (e.g. tabulating the study intervention characteristics and comparing against the planned groups for each synthesis (item #5)). | 7 |
|  | 13b | Describe any methods required to prepare the data for presentation or synthesis, such as handling of missing summary statistics, or data conversions. | 7 |
|  | 13c | Describe any methods used to tabulate or visually display results of individual studies and syntheses. | 7 |
|  | 13d | Describe any methods used to synthesize results and provide a rationale for the choice(s). If meta-analysis was performed, describe the model(s), method(s) to identify the presence and extent of statistical heterogeneity, and software package(s) used. | 7 |
|  | 13e | Describe any methods used to explore possible causes of heterogeneity among study results (e.g. subgroup analysis, meta-regression). | 7 |
|  | 13f | Describe any sensitivity analyses conducted to assess robustness of the synthesized results. | 7 |
| Reporting bias assessment | 14 | Describe any methods used to assess risk of bias due to missing results in a synthesis (arising from reporting biases). | N/A |
| Certainty assessment | 15 | Describe any methods used to assess certainty (or confidence) in the body of evidence for an outcome. | 7 |
| **RESULTS** | | |  |
| Study selection | 16a | Describe the results of the search and selection process, from the number of records identified in the search to the number of studies included in the review, ideally using a flow diagram. | 8 |
|  | 16b | Cite studies that might appear to meet the inclusion criteria, but which were excluded, and explain why they were excluded. | Figure 1 |
| Study characteristics | 17 | Cite each included study and present its characteristics. | 8 Table1 |
| Risk of bias in studies | 18 | Present assessments of risk of bias for each included study. | N/A |
| Results of individual studies | 19 | For all outcomes, present, for each study: (a) summary statistics for each group (where appropriate) and (b) an effect estimate and its precision (e.g. confidence/credible interval), ideally using structured tables or plots. | Table 2 |
| Results of syntheses | 20a | For each synthesis, briefly summarise the characteristics and risk of bias among contributing studies. | 8,9 |
|  | 20b | Present results of all statistical syntheses conducted. If meta-analysis was done, present for each the summary estimate and its precision (e.g. confidence/credible interval) and measures of statistical heterogeneity. If comparing groups, describe the direction of the effect. | 8,9 |
|  | 20c | Present results of all investigations of possible causes of heterogeneity among study results. | 8,9 |
|  | 20d | Present results of all sensitivity analyses conducted to assess the robustness of the synthesized results. | Supplementary Materials |
| Reporting biases | 21 | Present assessments of risk of bias due to missing results (arising from reporting biases) for each synthesis assessed. | N/A |
| Certainty of evidence | 22 | Present assessments of certainty (or confidence) in the body of evidence for each outcome assessed. | 8,9 |
| **DISCUSSION** | | |  |
| Discussion | 23a | Provide a general interpretation of the results in the context of other evidence. | 11 |
|  | 23b | Discuss any limitations of the evidence included in the review. | 14 |
|  | 23c | Discuss any limitations of the review processes used. | 14 |
|  | 23d | Discuss implications of the results for practice, policy, and future research. | 11,12 |
| **OTHER INFORMATION** | | |  |
| Registration and protocol | 24a | Provide registration information for the review, including register name and registration number, or state that the review was not registered. | 4 |
|  | 24b | Indicate where the review protocol can be accessed, or state that a protocol was not prepared. | 4 |
|  | 24c | Describe and explain any amendments to information provided at registration or in the protocol. | N/A |
| Support | 25 | Describe sources of financial or non-financial support for the review, and the role of the funders or sponsors in the review. | 15 |
| Competing interests | 26 | Declare any competing interests of review authors. | 15 |
| Availability of data, code and other materials | 27 | Report which of the following are publicly available and where they can be found: template data collection forms; data extracted from included studies; data used for all analyses; analytic code; any other materials used in the review. | N/A |

*From:* Page MJ, McKenzie JE, Bossuyt PM, Boutron I, Hoffmann TC, Mulrow CD, et al. The PRISMA 2020 statement: an updated guideline for reporting systematic reviews. BMJ 2021;372:n71. doi: 10.1136/bmj.n71

**Table S2** SWiM Reporting Checklist for systematic review of observational studies assessing generalizability of CVOTS

| **SWiM is intended to complement and be used as an extension to PRISMA** | | | |
| --- | --- | --- | --- |
| **SWiM reporting item** | **Item description** | **Page in manuscript where item is reported** | **Other*** |
| *Methods* | | | |
| **1** Grouping studies for synthesis | 1a) Provide a description of, and rationale for, the groups used in the synthesis (e.g., groupings of populations, interventions, outcomes, study design) | 7 |  |
|  | 1b) Detail and provide rationale for any changes made subsequent to the protocol in the groups used in the synthesis |  |  |
| **2** Describe the standardised metric and transformation methods used | Describe the standardised metric for each outcome. Explain why the metric(s) was chosen, and describe any methods used to transform the intervention effects, as reported in the study, to the standardised metric, citing any methodological guidance consulted | 7 |  |
| **3** Describe the synthesis methods | Describe and justify the methods used to synthesise the effects for each outcome when it was not possible to undertake a meta-analysis of effect estimates | 7 |  |
| **4** Criteria used to prioritise results for summary and synthesis | Where applicable, provide the criteria used, with supporting justification, to select the particular studies, or a particular study, for the main synthesis or to draw conclusions from the synthesis (e.g., based on study design, risk of bias assessments, directness in relation to the review question) | 5,6 |  |
| **SWiM reporting item** | **Item description** | **Page in manuscript where item is reported** | **Other*** |
| **5** Investigation of heterogeneity in reported effects | State the method(s) used to examine heterogeneity in reported effects when it was not possible to undertake a meta-analysis of effect estimates and its extensions to investigate heterogeneity | 6,7 |  |
| **6** Certainty of evidence | Describe the methods used to assess certainty of the synthesis findings | 7 |  |
| **7** Data presentation methods | Describe the graphical and tabular methods used to present the effects (e.g., tables, forest plots, harvest plots).  Specify key study characteristics (e.g., study design, risk of bias) used to order the studies, in the text and any tables or graphs, clearly referencing the studies included | 7 |  |
| *Results* | | | |
| **8** Reporting results | For each comparison and outcome, provide a description of the synthesised findings, and the certainty of the findings. Describe the result in language that is consistent with the question the synthesis addresses, and indicate which studies contribute to the synthesis | 8 |  |
| *Discussion* |  |  |  |
| **9** Limitations of the synthesis | Report the limitations of the synthesis methods used and/or the groupings used in the synthesis, and how these affect the conclusions that can be drawn in relation to the original review question | 14 |  |

PRISMA=Preferred Reporting Items for Systematic Reviews and Meta-Analyses.

*If the information is not provided in the systematic review, give details of where this information is available (e.g., protocol, other published papers (provide citation details), or website (provide the URL)).

The citation for the Synthesis Without Meta-analysis explanation and elaboration article is: Campbell M, McKenzie JE, Sowden A, Katikireddi SV, Brennan SE, Ellis S, Hartmann-Boyce J, Ryan R, Shepperd S, Thomas J, Welch V, Thomson H. Synthesis without meta-analysis (SWiM) in systematic reviews: reporting guideline BMJ 2020;368:l6890 <http://dx.doi.org/10.1136/bmj.l6890>

**Table S3** Search strategy for systematic review of observational studies assessing generalizability of CVOTS using OVID MEDLINE database

|  | **Search Entry** |
| --- | --- |
| 1. | Observational study.pt. or cohort studies/ or case-control studies/ or cross-sectional studies/ or (real-world or observation* or nonrandomi#ed or non randomi#ed or case control or cohort stud* or cross sectional or (emulat* adj3 trial?)).mp. \| |
| 2. | Randomized Controlled Trial/ or Randomized Controlled Trials as Topic/ or Clinical Trial/ or Clinical Trials as Topic/ or trial?.mp. |
| 3. | (major adverse cardiovascular event* or (cardiovascular adj3 outcome*) or all cause mortality).mp. \| |
| 4. | Diabetes Mellitus/ or Diabetes Mellitus, Type 2/ or (antidiabet* or diabet* or t2d or t2dm).mp. |
| 5. | (emul* or compar* or applicab* or generali#ab* or transportability).mp. |
| 6. | 1 AND 2 AND 3 AND 4 AND 5 |

**Table S4** Search strategy for systematic review of observational studies assessing generalizability of CVOTS using OVID EMBASE + EMBASE CLASSIC database

|  | **Search Strategy** |
| --- | --- |
| 1. | Observational study.pt. or cohort analysis/ or case control study/ or cross-sectional study/ or (real-world or observation* or nonrandomi#ed or non randomi#ed or case control or cohort stud* or cross sectional or (emulat* adj3 trial?)).mp. |
| 2. | Randomized Controlled Trial/ or Clinical Trial/ or trial?.mp. |
| 3. | \| (major adverse cardiovascular event* or (cardiovascular adj3 outcome*) or all cause mortality).mp. |
| 4. | Diabetes Mellitus/ or non insulin dependent diabetes mellitus/ or (antidiabet* or diabet* or t2d or t2dm).mp. |
| 5. | (emul* or compar* or applicab* or generali#ab* or transportability).mp. |
| 6. | 1 AND 2 AND 3 AND 4 AND 5 |

**Table S5** Search strategy for systematic review of observational studies assessing generalizability of CVOTS using Cochrane CENTRAL database

|  | **Search Strategy** |
| --- | --- |
| 1. | "observational study":pt OR [mh ^"cohort studies"] OR [mh ^"case-control studies"] OR [mh ^"cross-sectional studies"] OR (real-world OR observation* OR nonrandomi#ed OR "non randomi") |
| 2. | [mh ^"Randomized Controlled Trial"] OR [mh ^"Randomized Controlled Trials as Topic"] OR [mh ^"Clinical Trial"] OR [mh ^"Clinical Trials as Topic"] OR trial?:ti,ab,kw |
| 3. | (("major adverse cardiovascular" NEAR/2 event*) OR (cardiovascular NEAR/3 outcome*) OR "all cause mortality"):ti,ab,kw |
| 4. | [mh ^"Diabetes Mellitus"] OR [mh ^"Diabetes Mellitus, Type 2"] OR (antidiabet* OR diabet* OR t2d OR t2dm ):ti,ab,kw |
| 5. | (emul* OR compar* OR applicab* OR generali#ab* OR transportability):ti,ab,kw |
| 6. | 1 AND 2 AND 3 AND 4 AND 5 |

**Table S6** A list of completed cardiovascular outcome trials and their drug class and molecule

| Drug Class | Trial Name | Drug Molecule |
| --- | --- | --- |
| DPP-4 Inhibitor | SAVOR-TIMI^41^ | Saxaglibptin |
|  | EXAMINE | Alogliptin |
|  | TECOS^42^ | Sitagliptin |
|  | CAROLINA^51^ | Lingaliptin |
|  | CARMELINA^40^ | Linagliptin |
|  | MK-3102^52^ | Omarigliptin |
| GLP-1 receptor agonist | ELIXA^44^ | Lixisenatide |
|  | LEADER^36^ | Liraglutide |
|  | SUSTAIN-6^39^ | Semaglutide |
|  | PIONEER-6^37^ | Semaglutide |
|  | EXSCEL^35^ | Exenatide-pragmatic trial |
|  | REWIND^38^ | Dulaglutide |
|  | FREEDOM-CVO^45^ | Exenatide in DUROS |
|  | HARMONY^46^ | Albiglutide |
| SGLT-2 inhibitor | EMPA-REG^32^ | Empagliflozin |
|  | CANVAS^33^ | Canagliflozin |
|  | DECLARE^34^ | Dapagliflozin |
|  | VERTIS-CV^43^ | Ertugliflozin |

**Table S7** Standardized differences computed for observational study compared to RCT hazard ratios

| Study | RCT | Sample Size | |  |  | RCT | |  |
| --- | --- | --- | --- | --- | --- | --- | --- | --- |
|  |  | Exposure | Comparator | HR | 95% CI | HR | 95% CI | SD |
| Sciannameo, 2021 | EMPA-REG^32^ | N/A | N/A | 0.88 | 0.74, 1.03 | 0.86 | 0.74, 0.99 | 0.20 |
| Sciannameo, 2021 | DECLARE^34^ | N/A | N/A | 0.94 | 0.84, 1.04 | 0.93 | 0.84, 1.03 | 0.14 |
| Sciannameo, 2021 | EXSCEL^35^ | N/A | N/A | 0.92 | 0.82, 1.02 | 0.91 | 0.83, 1.00 | 0.15 |
| Abrahami, 2020 | LEADER^36^ | 1 868 | 25 895 | 1.03 | 0.82, 1.30 | 0.87 | 0.78, 0.97 | 1.09 |
| Abrahami, 2020 | LEADER^36^ | 1 864 | 32 899 | 0.97 | 0.78, 1.22 | 0.87 | 0.78, 0.97 | 0.73 |
| Franklin, 2020 | LEADER^36^ | 4 668 | 4 672 | 0.82 | 0.76, 0.87 | 0.87 | 0.78, 0.97 | -0.88 |
| Sciannameo, 2021 | LEADER^36^ | N/A | N/A | 0.88 | 0.77, 0.99 | 0.87 | 0.78, 0.97 | 0.13 |
| Sciannameo, 2021 | PIONEER-6^37^ | N/A | N/A | 0.76 | 0.41, 1.10 | 0.79 | 0.57, 1.11 | -0.13 |
| Sciannameo, 2021 | REWIND^38^ | N/A | N/A | 0.87 | 0.76, 0.98 | 0.88 | 0.79, 0.99 | -0.12 |
| Sciannameo, 2021 | SUSTAIN-6^39^ | N/A | N/A | 0.73 | 0.47, 0.99 | 0.74 | 0.58, 0.95 | -0.06 |
| Sciannameo, 2021 | SAVOR-TIMI^41^ | N/A | N/A | 0.99 | 0.87, 1.10 | 1.00 | 0.89, 1.12 | -0.12 |
| Sciannameo, 2021 | DECLARE^34^-HHF/CV Death | N/A | N/A | 0.86 | 0.73, 0.99 | 0.83 | 0.73, 0.95 | 0.30 |

Abbreviations: RCT: Randomized Controlled Trials; HR: Hazard Ratio; CI: Confidence Interval; SD: Standardized Difference; HHF: Hospitalization due to heart failure; CV: Cardiovascular

**Figure S1** Distribution of percent eligible for inclusion in CVOT of SGLT-2 inhibitors in real-world populations by mean age (i) and by sex (ii)

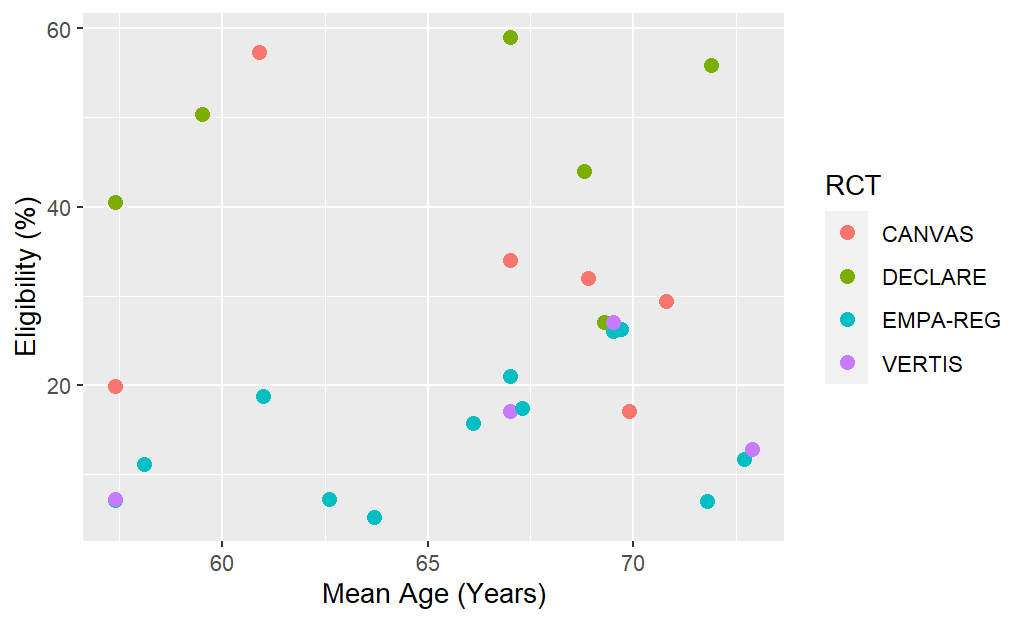

i.

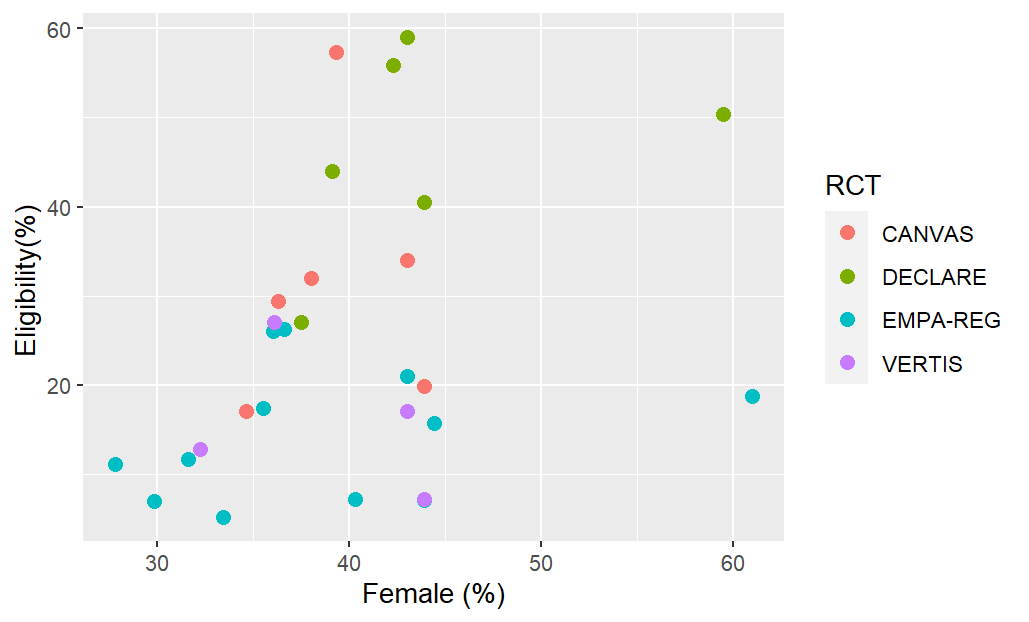

ii.

**Figure S2** Distribution of percent eligible for inclusion in CVOT of GLP-1 Receptor Agonists in real-world populations by mean age (i) and by sex (ii)

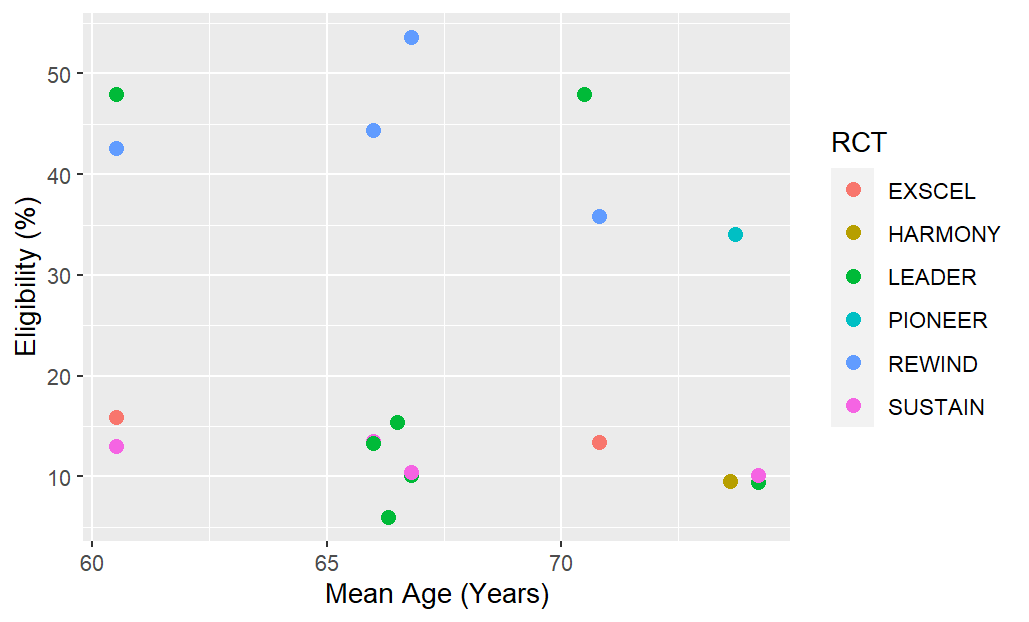

i.

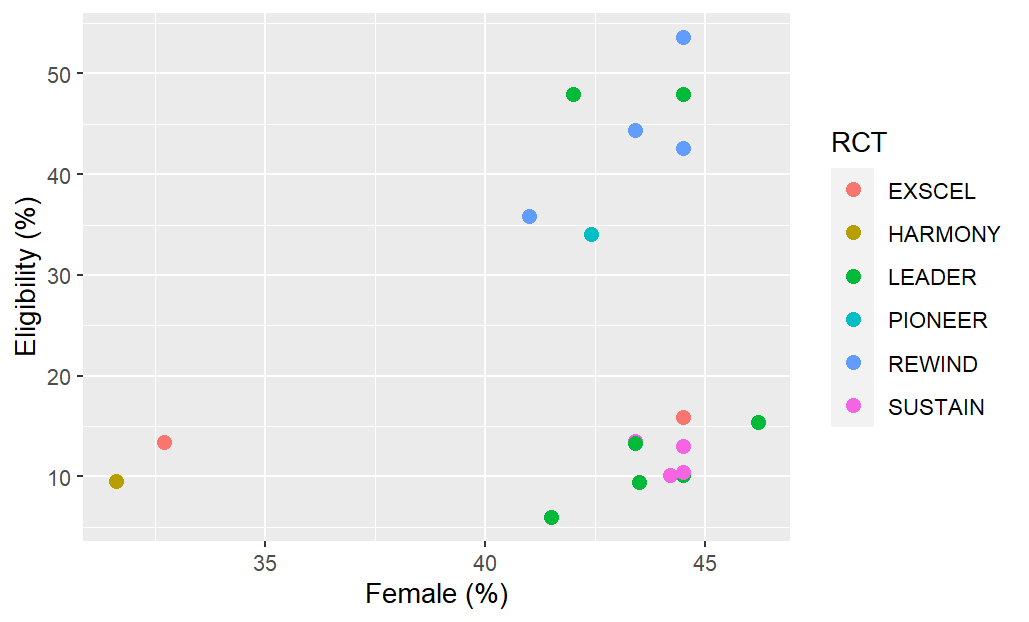

ii

**Figure S3** Standardized Mean Differences of patient characteristics between GLP-1 Receptor Agonist CVOTs and the study population from the RWD study from Sciannameo, 2020.

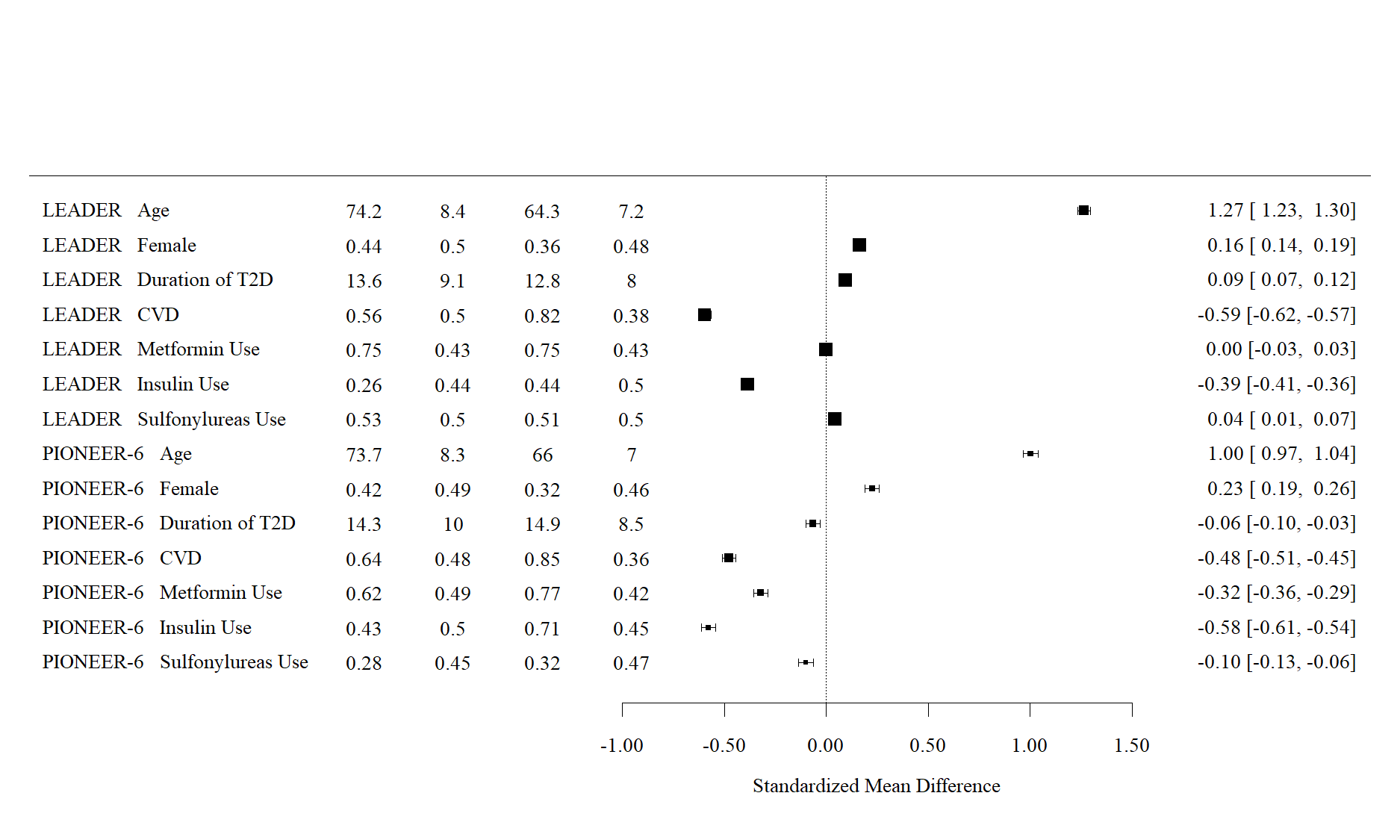

**Study	Characteristics	RWD	RCT**

**Mean/prop	SD	Mean/Prop	SD**
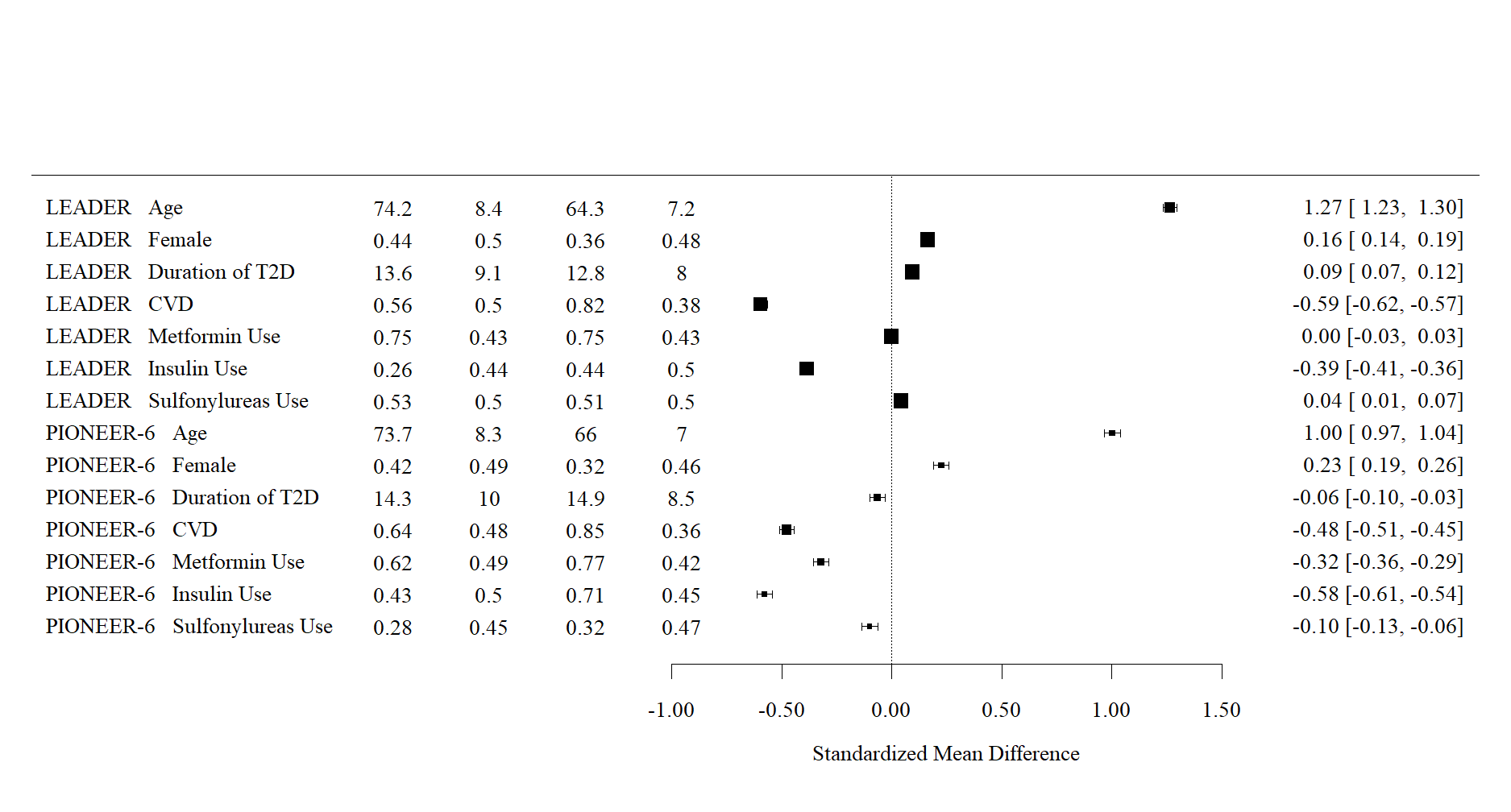
 **SD	Mean/%	SD**

**SMD [95% CI]**

Abbreviations: CVD: cardiovascular disease, CVOT: cardiovascular outcome trial, Prop: Proportion; RCT: randomized controlled trial, RWE: real-world evidence, SD: standard deviation, SMD: standardized mean difference, T2D: Type 2 diabetes,

**Figure S4** Select standardized mean difference of patient characteristics for CVOT and RWD for the DECLARE trial

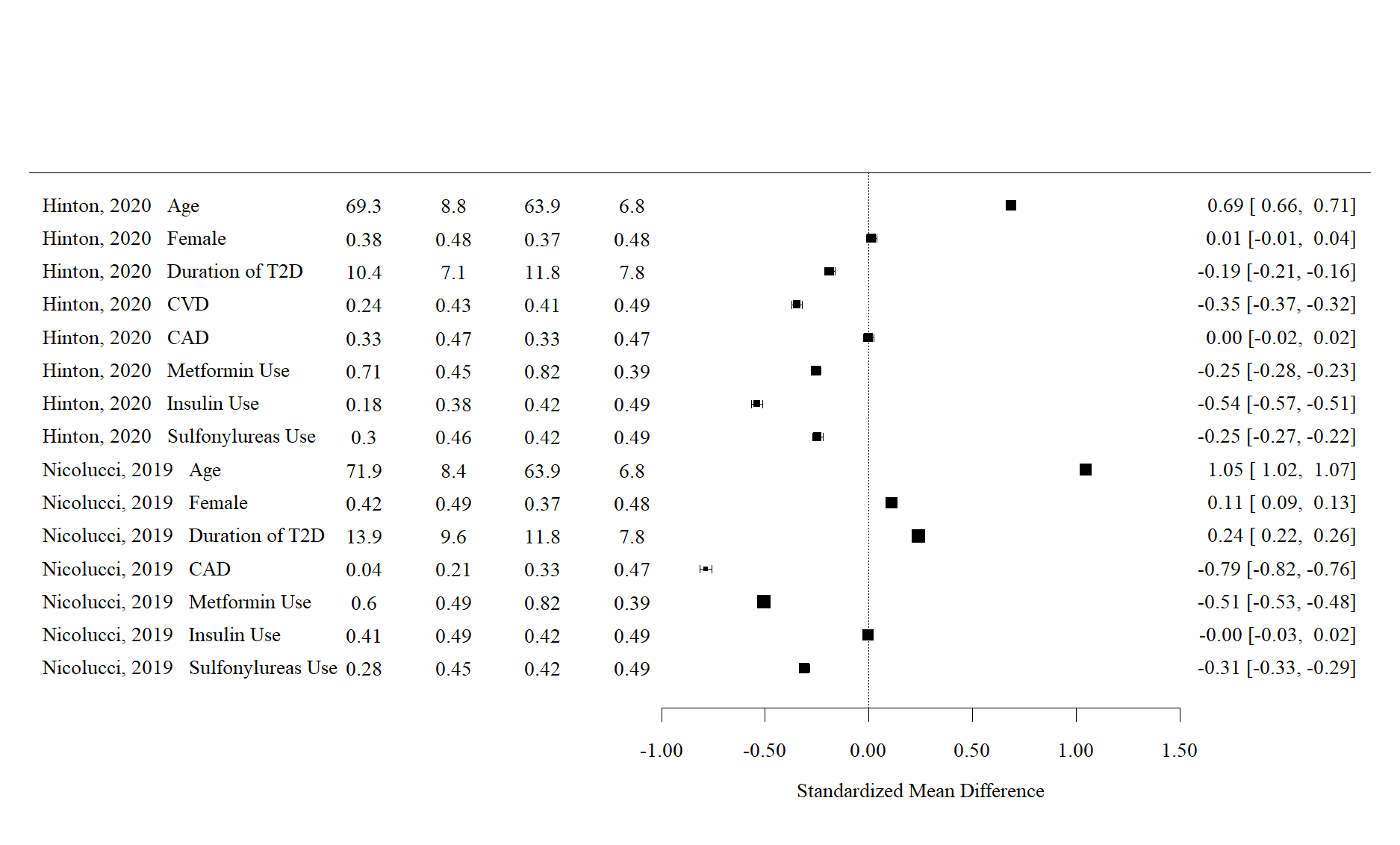

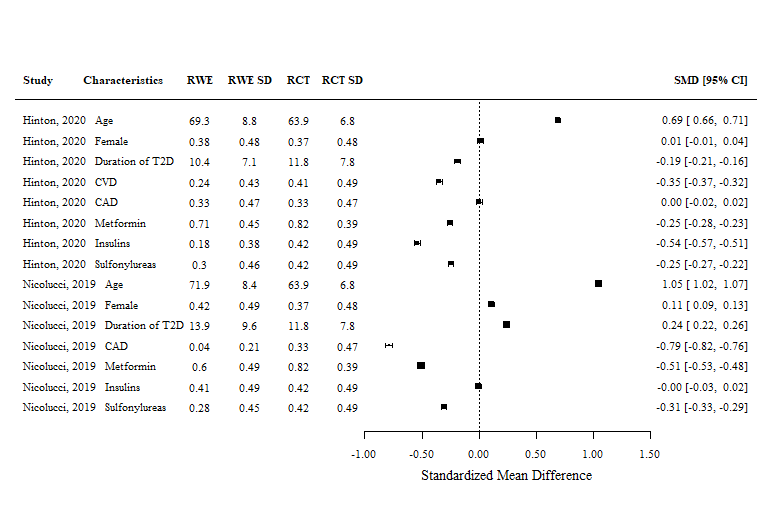

**SMD [95% CI]**

**Study	Characteristics	RWD	RCT**

**Mean/prop	SD	Mean/Prop	SD**
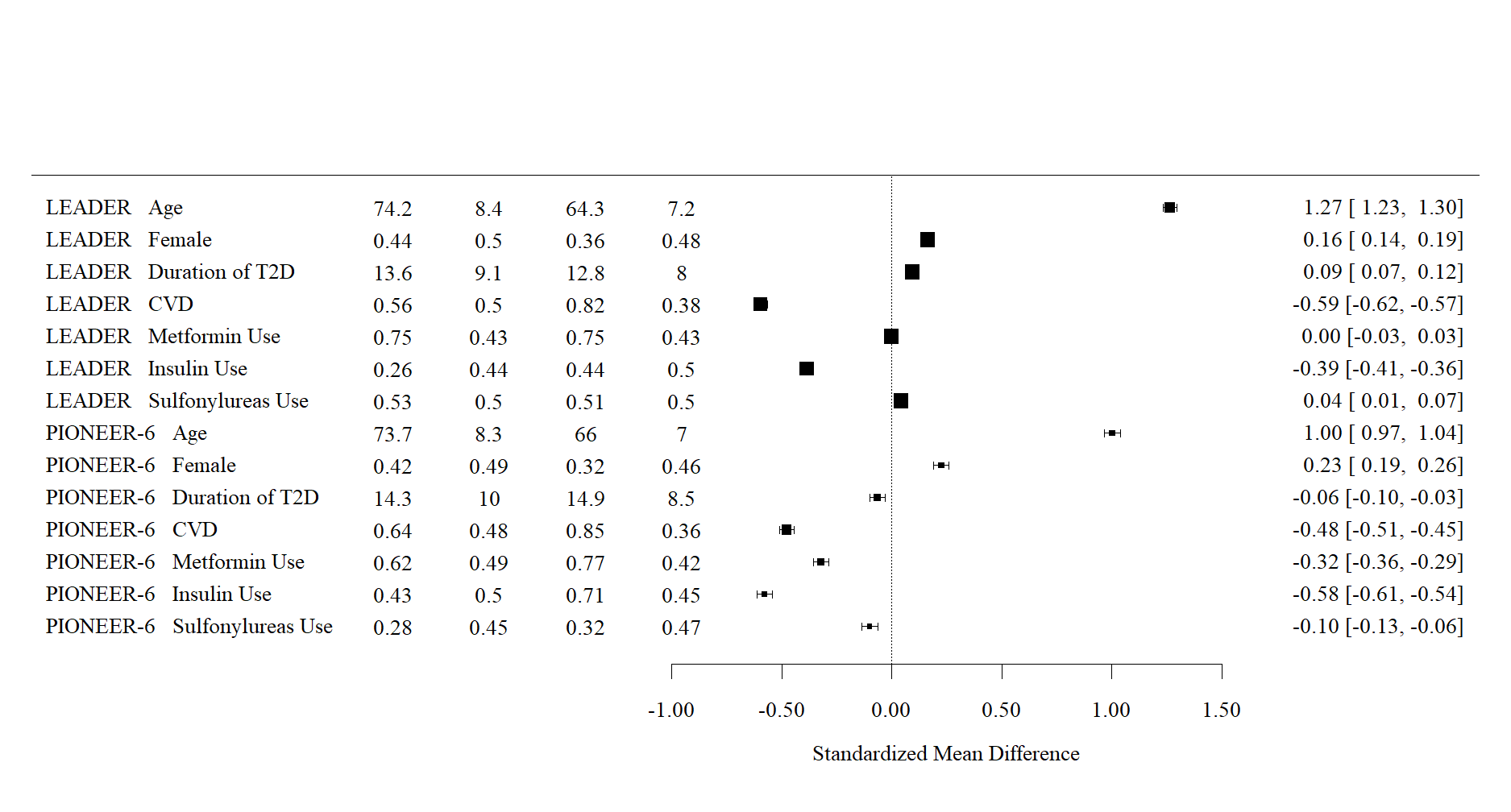
 **SD	Mean/%	SD**

Abbreviations: CAD: coronary artery disease, CVD: cardiovascular disease, CVOT: cardiovascular outcome trial, Prop: Proportion, RCT: randomized controlled, RWE: real-world evidence, RWD: real-world data, SD: standard deviation, trial, SMD: standardized mean difference, T2D: Type 2 diabetes

**Figure S5** Select standardized mean difference of patient characteristics for CVOT and RWD for CANVAS trial

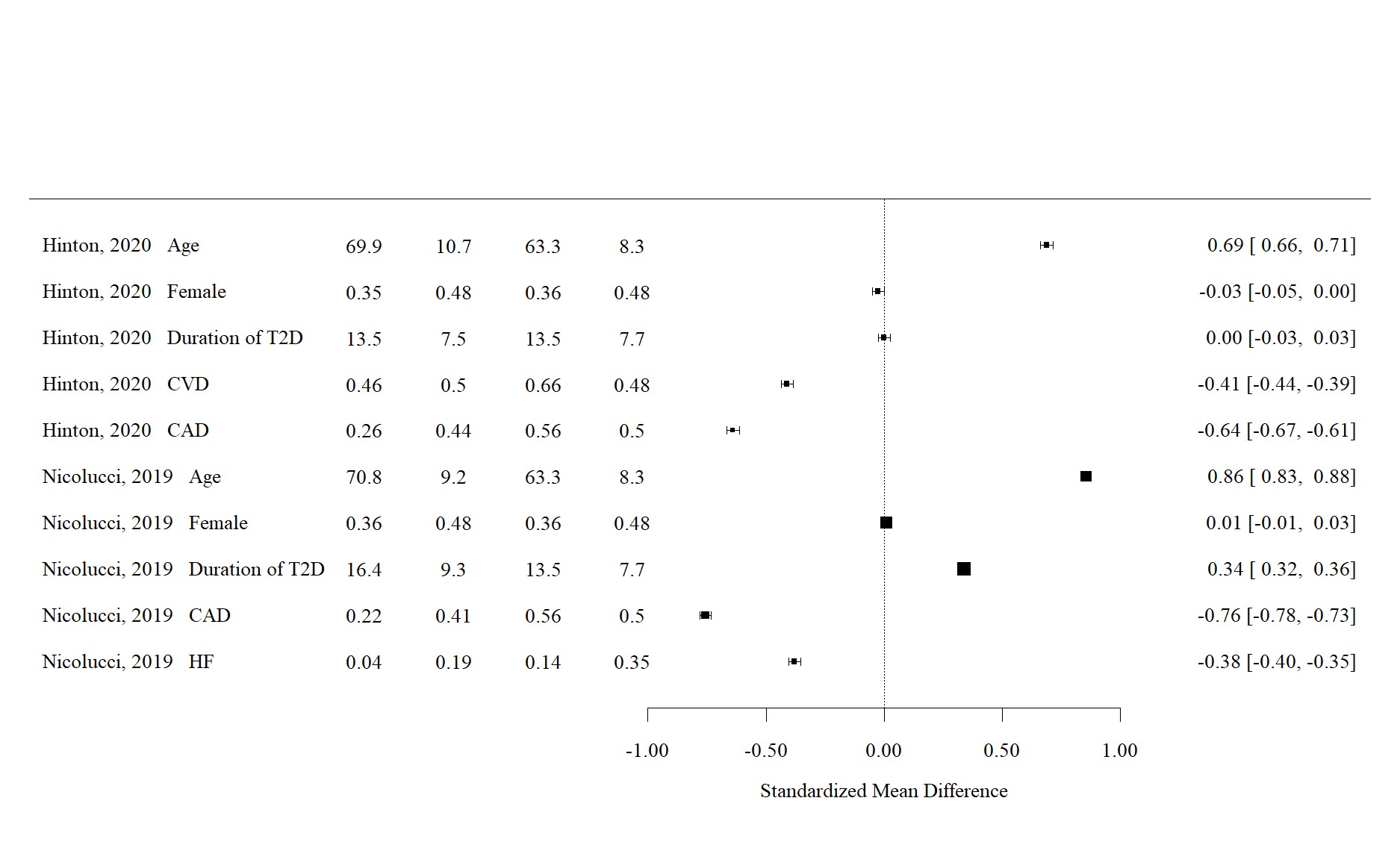

**SMD [95% CI]**

**Study	Characteristics	RWD	RCT**

**Mean/Prop	SD	Mean/Prop	SD**
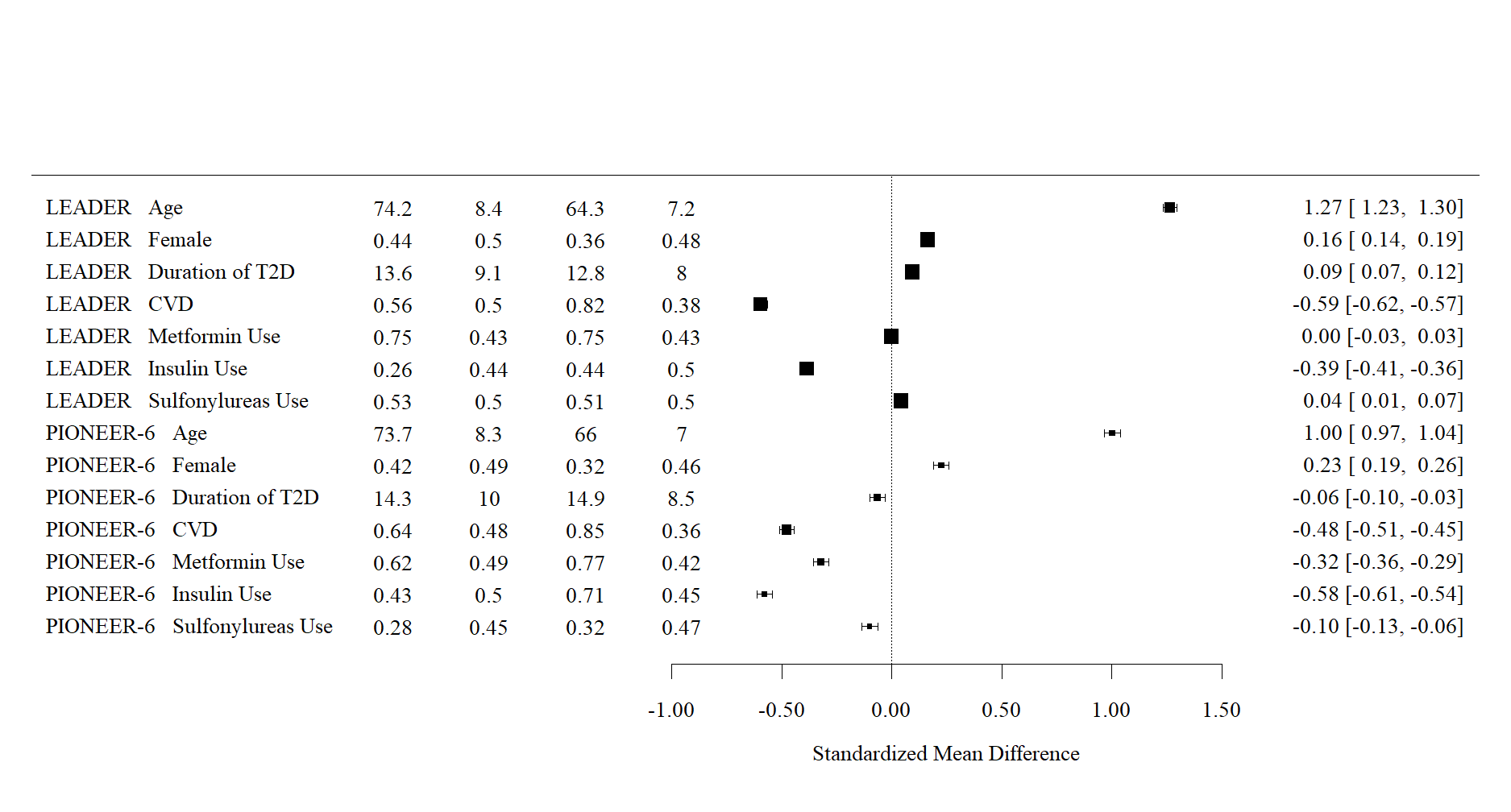
 **SD	Mean/%	SD**

Abbreviations: CAD: coronary artery disease, CVD: cardiovascular disease, CVOT: cardiovascular outcome trial, HF: heart failure, RCT: randomized controlled, RWE: real-world evidence, RWD: real-world data, SD: standard deviation, trial, SMD: standardized mean difference, T2D: Type 2 diabetes
